# Citywide indoor air sampling mirrors wastewater and clinical case surveillance of respiratory viruses

**DOI:** 10.1101/2025.10.13.25337283

**Authors:** Hannah J Barbian, Erin P Newcomer, Sofiya Bobrovska, Rachel Poretsky, Stephanie Greenwald, Sarah M Owens, Anuj Tiwari, Rachel J Berkowitz, Samantha Smith, Dorothy Foulkes, Stefan J Green, Dolores Sanchez Gonzales, Chi-Yu Lin, Adam Horton, Modou Lamin Jarju, Rosemarie Wilton, Mary K Hayden, Stephanie R Black, V Eloesa McSorely, Alyse Kittner

## Abstract

Wastewater surveillance of respiratory pathogens can provide timely estimates of viral activity and disease trends in a population. Indoor air surveillance could be used similarly with some advantages but remains largely unvalidated at the community-scale. Here, an indoor air surveillance program was employed as part of public health environmental surveillance in Chicago, Illinois, USA. Ten air samplers were placed in healthcare and congregate living settings across the city. Weekly air samples were evaluated for influenza A, influenza B, respiratory syncytial virus, and SARS-CoV-2 over two respiratory virus seasons. Citywide, aggregated air sample positivity and viral load were closely correlated with local clinical case and wastewater surveillance data across all respiratory viruses. Virus trends in air data often preceded clinical and wastewater, although this varied across pathogens and respiratory virus seasons. Further, whole-genome sequencing of SARS-CoV-2 showed close correlation of variant proportions across all datasets. At the building-scale, air samples obtained from a single sampling device provided efficient respiratory virus surveillance, with well-correlated estimates of respiratory pathogens. These data demonstrate that air surveillance can provide accurate estimates of respiratory virus infections and variants at a building or community-scale, serving as an alternative or complementary tool for public health environmental surveillance.

## INTRODUCTION

Over the past five years, environmental surveillance has emerged as an essential tool in public health for monitoring community-level infectious disease, providing an accurate indicator of local pathogen activity complementary to traditional case-based surveillance. Compared to traditional clinical case-based surveillance, environmental surveillance does not require testing of symptomatic individuals, reducing the bias of test-seeking behavior as well as the lag time from disease transmission to detection by surveillance methods, sometimes resulting in an early indicator of disease activity^1–3^. Because of this, integration of wastewater data into public health programs has rapidly expanded following its initial development in response to the COVID-19 pandemic^4^. Public health jurisdictions use wastewater surveillance data to anticipate local peaks in seasonal viruses, detect emerging or rare pathogens, and inform public health policies^5^. The scope of wastewater surveillance has since expanded beyond SARS-CoV-2 to include other pathogens of public health concern such as influenza (including H5), respiratory syncytial virus (RSV), and mpox^6–9^. Further, genomic surveillance has become an informative component of wastewater programs, enabling the detection and characterization of pathogen strains and variants circulating within populations. SARS-CoV-2 genomic surveillance from wastewater samples has provided key insights into the emergence and spread of variants of concern, often preceding clinical detection^10^.

Like wastewater surveillance, indoor air surveillance can be used for environmental detection of infectious disease activity, capturing both symptomatic and asymptomatic shedding^11^. While indoor air samples have historically faced sensitivity issues associated with low biomass specimens, recent studies suggest that respiratory pathogens can be readily detected in air samples from high-risk indoor settings such as hospitals and schools^11–14^. Further, it has been shown that respiratory pathogen levels in air samples from medical settings reflects case-based surveillance^6,14,15^. Air surveillance may be able to fill some gaps that are difficult to achieve using wastewater sampling. Air samplers are mobile, allowing them to be applied according to surveillance need and easily target highly specific populations. Wastewater data at the sewer or treatment plant level serving large populations can dilute signals of rare or emerging pathogens and preclude attributing pathogen signals to specific locations or subpopulations, limiting their use for targeted interventions.^16^ Air surveillance is easier to deploy at individual buildings, with placement of an air sampler taking only a few minutes and not requiring coordination with building engineers or requiring confirmatory dye tests, allowing rapid response in outbreak situations affecting specific locations or populations. Air samples may better reflect building occupants compared to wastewater, as not everyone use the restroom in buildings they transiently occupy (e.g. schools, outpatient medical facilities, workplaces, nurseries with diapered individuals). While air samplers surveille smaller populations than sewer or treatment plant wastewater samples, possibly limiting their sensitivity and representativeness, aggregating samples or results from a network of several air samplers may overcome this limitation. Finally, some respiratory pathogens may not be shed into wastewater at concentrations high enough for detection or characterization; environmental air samples may be a preferred sample source for some pathogens. Despite these promising qualities, air surveillance has so far not been validated at a community-wide scale for public health infectious disease surveillance.

Here, we aimed to deploy and evaluate a citywide indoor air sampling program as a novel mechanism for public health environmental infectious disease surveillance. Air samplers were placed at various medical and congregate living locations around the City of Chicago by the Chicago Department of Public Health (CDPH). Results of air sample surveillance were aggregated and compared to established community surveillance mechanisms: traditional clinical case-based surveillance and local wastewater surveillance. Influenza A, influenza B, RSV, and SARS-CoV-2 were selected as they are i) pathogens of public health concern, ii) respiratory viruses with temporal variation, and iii) also targeted by local case and wastewater surveillance. Further, one congregate living facility participated in both building air and wastewater surveillance; this site was assessed as an example of facility-level environmental surveillance. As public health programs continue to diversify environmental surveillance strategies, air sampling could offer a valuable new tool for early detection, disease forecasting, and outbreak mitigation if able to reflect local infectious disease activity.

## METHODS

### Air sample collection

AerosolSense samplers (ThermoFisher) were deployed at 10 locations and placed on a table at each site (approximately 0.75 meters above the floor) (Table 1, Figure 1a-b). AerosolSense cartridges (ThermoFisher) were inserted into the sampler and collected at the instrument default of 200 L/min for 7 continuous days (Figure 1b). At the end of the sampling week, the cartridges were removed and new cartridges were immediately inserted for the subsequent week. Air sample cartridges were transported to the Regional Innovative Public Health Laboratory (RIPHL), an academic partner of CDPH at Rush University Medical Center, at room temperature via local courier. Cartridge collection dates and internal IDs were recorded using QR code stickers applied to each cartridge which were associated with an electronic record in web-based database REDCap^17^. Facility staff at each site were responsible for weekly cartridge change on Mondays.

**Figure 1.**
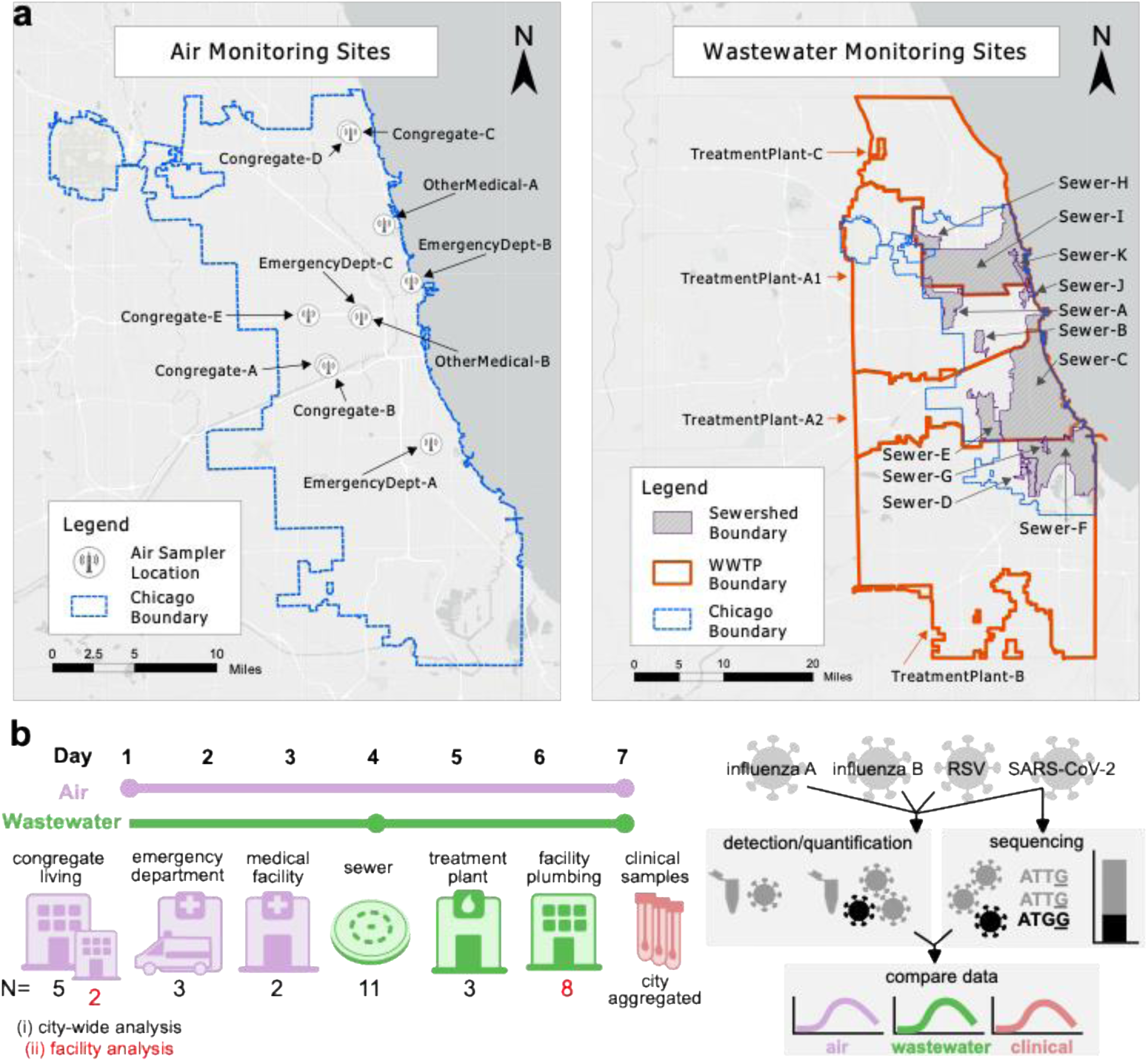
Study overview and sampling sites. a) Map of air and wastewater sampling sites. b) Study overview showing air (purple) and wastewater (green) weekly sampling strategies (circle indicates sampling start/end); types and numbers of air (purple) and wastewater (green) sampling sites contributing to (black numbers) and single-facility (red numbers) analyses); and viruses targeted for analyses.

**Table 1.** Air sampling locations.

| Site ID | Site description | Placement description | Room occupancy* |
| --- | --- | --- | --- |
| Congregate A | Congregate living facility with transient adult population | Waiting room seating area | 20-30 |
| Congregate B | Congregate living facility with transient adult population | Waiting room seating area | 30 |
| Congregate C | Congregate living facility with consistent population of children and adults | Recreation room | 80-100 |
| Congregate D | Congregate living facility with consistent population of children and adults | Seating area attached to dining area | 90-110 |
| Congregate E | Long term acute care hospital | Non-clinical hallway | 200-300 |
| Emergency Dept A | Emergency department within a children's acute care hospital | Clinical hallway | unavailable |
| Emergency Dept B | Emergency department within<br>a free-standing children's<br>acute care hospital | Waiting room seating<br>area | 250-350 |
| Emergency Dept C | Emergency department within<br>an acute care hospital serving<br>adults and children | Waiting room entrance /<br>reception desk | 200-300 |
| Other Medical A | Specialty clinic serving adults | Waiting room entrance /<br>reception desk | 20-25 |
| Other Medical B | Acute care hospital serving<br>adults and children | Dining area and lounge | 250-350 |

\*Estimated occupancy per day

### Wastewater collection

Chicago wastewater surveillance began in 2020 as a partnership between CDPH, local utilities, and academic, municipal, and industry partners. The current program primarily focuses on community-level surveillance from neighborhood sewers and pumping stations. However, facility-level testing was implemented at several settings of public health importance during the pandemic to support outbreak detection and response. The Illinois Department of Public Health (IDPH) samples wastewater treatment plants which serve both Chicago and parts of the surrounding Cook County; data from these samples are publicly available (https://iwss.uillinois.edu/). For this work, duplicate 50 ml wastewater samples were collected from locations across the Chicagoland area including sewers, facilities, and treatment plants (Table 2). Sewers and facilities employed passive sampling (Moore swabs consisting of sterile cotton gauze suspended in maintenance holes) while treatment plants provided time-proportional 24-hour flow-weighted composite samples. Samples were transported to the University of Illinois Chicago on ice.

**Table 2.** Wastewater sampling locations.

| **Site_ID** | **Population**<br>**Estimate** | **Access point**<br>**description** | **Sample type** |
| --- | --- | --- | --- |
| **Sewer A** | 83,455 | Community manhole | Moore swab |
| **Sewer B** | 29,370 | Community manhole | Moore swab |
| **Sewer C** | 467,536 | Pumping Station | Moore swab |
| **Sewer D** | 23,475 | Community manhole | Moore swab |
| **Sewer E** | 77,799 | Community manhole | Moore swab |
| **Sewer F** | 125,995 | Pumping Station | Moore swab |
| **Sewer G** | 3,816 | Community manhole | Moore swab |
| **Sewer H** | 23,324 | Community manhole | Moore swab |
| **Sewer I** | 721,207 | Pumping Station | Moore swab |
| **Sewer J** | 24,099 | Community manhole | Moore swab |
| **Sewer K** | 54,362 | Community manhole | Moore swab |
| **Treatment Plant**<br>**A1** | 1,127,736 | Wastewater Treatment<br>Plant, Inlet 1 | Autosampler |
| **Treatment Plant**<br>**A2** | 1,127,736 | Wastewater Treatment<br>Plant, Inlet 2 | Autosampler |
| **Treatment Plant**<br>**B** | 1,134,897 | Wastewater Treatment<br>Plant | Autosampler |
| **Treatment Plant**<br>**C** | 1,127,736 | Wastewater Treatment<br>Plant | Autosampler |
| **Site A** | 686 | Facility housing adults | Moore swab |
| **Site B** | 987 | Facility housing adults | Moore swab |
| **Site C** | 200 | Facility housing adults | Moore swab |
| **Site D** | 963 | Facility housing adults | Moore swab |
| **Site E** | 434 | Facility housing adults | Moore swab |
| **Site F** | 976 | Facility housing adults | Moore swab |
| **Site G** | 1577* | Facility housing adults | Moore swab |
| **Site H** | 1577* | Facility housing adults | Moore swab |

\*Multiple sewer access points (Site G and Site H) for a single building. Estimated population represents entire building

### Detection of respiratory pathogens from air

Air cartridges were stored at 4°C upon arrival at RIPHL and were processed within four days of arrival as previously described^14^. Briefly, a biosafety cabinet used exclusively for low-biomass pre-PCR specimens was cleaned using UV light, 70% ethanol and RNase AWAY (Applied Biological Materials). Inside the cabinet, both membranes were removed from each cartridge using a disposable sterile forceps and transferred to separate microcentrifuge tubes containing 500 µl DNA/RNA Shield Stabilization Solution (Zymo Research). One membrane was immediately processed, and the other was stored at −80°C. Processed membranes were manually depressed with a pipette tip until fully suspended then 300 µl was subjected to nucleic acid extraction using the Maxwell RSC Viral Total Nucleic Acid Purification Kit and Maxwell RSC automated extraction instrument (Promega) according to manufacturer instructions. An unused cartridge was included as an extraction blank in every run. Nucleic acid extracts were eluted in 75 µl nuclease-free water.

Total nucleic acid extracts were tested via quantitative PCR (qPCR) within 24 hours of extraction for respiratory viruses as previously described^14^. Briefly, pathogen-specific RSV A/B, influenza A, influenza B, and SARS-CoV-2 assays in TrueMark custom plates (ThermoFisher) were run according to manufacturer instructions. An internal control assay for human RNaseP was multiplexed in each well. 5 µl air sample extract was added to each reaction and plates were run on a Quantstudio 5 real-time PCR instrument with standard cycling mode (ThermoFisher). Constant baseline and threshold values were used to generate cycle threshold (Ct) values across all runs using QuantStudio software. Raw data were imported into RStudio for quality control filtering and interpretation^18^. Quality controls included confirming the extraction blank was negative across all pathogens and within-sample RNase P Ct values across pathogen wells were below standard deviation 1.5. Samples not meeting these were repeated or excluded. Virus detection was assigned “positive” if the Ct value was below the cutoff Ct determined through limit of detection studies, described previously, and assigned “negative” otherwise^14^.

### Detection of respiratory pathogens from wastewater

The wastewater sample processing method was adapted from Karthikeyan et al. (2021)^1^. Briefly, once samples arrived in the lab, 10 mL of wastewater were spiked with 80 µL of Bovine Coronavirus (BCoV) solution as a process control, all handled in a biosafety cabinet cleaned with UV light, 70% ethanol and RNase AWAY (Applied Biological Materials). Wastewater was then concentrated on the KingFisher Apex Purification System (ThermoFisher Scientific) using magnetic Nanotrap Microbiome A Particles (Ceres Nanosciences). Finally, total nucleic acids were extracted on the KingFisher Apex using the MagMax Wastewater Ultra Nucleic Acid Isolation Kit (ThermoFisher Scientific).

Pathogen targets SARS-CoV-2, influenza A and B, and RSV A/B detection used commercial kits according to the manufacturer’s instructions for respiratory pathogens (GT-Digital Flu;SC2;RSV Wastewater Surveillance Panel for the QIAcuity™ Digital PCR System, GT Molecular, Fort Collins, CO). For dPCR, 7.6 µL of molecular grade water and 5 µL extracted RNA diluted 1:4 with molecular grade water for a total volume of 20 µL per sample. dPCR was done on a QIAcuity instrument in 26k 24-well nanoplates (Qiagen). All dPCR was reported as gene copies per liter (copies/L) of starting wastewater. Quality controls included method blanks, negative, and positive controls. Raw wastewater concentrations were compared against the respective limits of detection (LOD), determined through serial dilutions of positive controls.

### Sequencing of SARS-CoV-2 from air

SARS-CoV-2 libraries were prepared from air sample total nucleic acid extracts within 2 weeks of sample collection as previously described^14^. Briefly, QIAseq DIRECT SARS-CoV-2 kits (QIAGEN) were used according to manufacturer instructions. Libraries were sequenced on an Illumina NovaSeq X sequencer, targeting 5 million clusters per sample (2×150 base reads). Raw reads were subsampled to a maximum of 20 million paired-end reads. Subsampled reads were analyzed using the Freyja_FastQ_PHB pipeline v2.3.0^19^ using the barcodes published on 3-12-2025, which calls Freyja^10^ for depth-weighted demixing of SARS-CoV-2 variants. Samples with <40% coverage were excluded from analysis. Air sample sequences were uploaded to NCBI sequence read archive (SRA), BioProject PRJNA1337702.

### Sequencing of SARS-CoV-2 from wastewater

Qiagen’s QIAseq DIRECT SARS-CoV-2 kit was used to generate targeted whole-genome SARS-CoV-2 libraries using Qiagen’s Booster primers using the manufacturer’s protocol. Library concentrations were quantified on the Tecan Infinite M200 Pro using PicoGreen dye (Invitrogen). Libraries were then pooled equimolar with an Opentrons OT2 liquid handling robot. The final pool was quantified using a Qubit 4 Fluorometer. Libraries were sequenced on an Illumina NextSeq 2000 on a 2×151bp run using P1 Reagents (300 Cycles) targeting an average of 50,000 reads per sample. Wastewater sequences have been uploaded to NCBI BioProject PRJNA989260. Sample collection source for wastewater treatment plant samples were provided by internal communication with IDPH.

To assess variant proportions, wastewater fastqs were aligned to the Wuhan-Hu-1 reference (MN908947.3) using Minimap 2^20^. After alignment, resulting BAM files were filtered, sorted, and indexed using SAMtools ^21^. iVar was then used to clip the primers using a BED file for the Qiagen DIRECT SARS-CoV-2 Kit^22^. Resorted clipped BAM files were then used as input into Freyja using the barcodes published on 3-12-2025^10^. All samples with >40% genome coverage were retained.

### Sequencing of SARS-CoV-2 from clinical specimens

CDPH recruited a random convenience sample of remnant SARS-CoV-2 positive nasal swab specimens from several Chicago acute-care hospitals for baseline genomic surveillance^23^. Remnant specimens were shipped weekly by local courier service to RIPHL. Specimens were heat inactivated upon arrival then stored at 4°C until they were processed in weekly or bi-weekly batches. Total nucleic acids were extracted using a Maxwell RSC and Maxwell RSC Viral Total Nucleic Acid Purification Kit. Extracted samples underwent reverse transcription using High Capacity Reverse Transcription Kit (Applied Biosystems). Libraries were prepared according to manufacturer instructions using QIAseq DIRECT SARS-CoV-2 kits (QIAGEN) for samples processed before May 2024 and COVIDseq Assay kits (Illumina) for subsequent samples.

Raw reads were subsampled to a maximum of 2 million paired-end reads. Subsampled reads were analyzed using the TheiaCoV_Illumina_PE_PHB pipeline v2.2.0 with minimum read depth of 25^19^. SARS-CoV-2 lineage designation and quality assessment is performed on consensus sequences using the most up-to-date versions of pangolin pdata and NextClade dataset tags^24,25^. Samples were retained for analysis if they met the following quality requirements: <3000 N, good or mediocre Nextclade quality status, and Pango lineage assigned. Sample lineage assignments are updated and maintained in TheiaGenEpi.org and were downloaded on 2025-04-09. Consensus sequences were uploaded to NCBI Virus, BioProject PRJNA903175.

### Analysis of SARS-CoV-2 variants across air, wastewater, and clinical samples

Variants were aggregated into parent variant groups based on variants delineated by the CDC Variants and Genomic Surveillance page^26^ that reached above 10% relative abundance in clinical samples for at least one collection week, with weeks corresponding to Morbidity and Mortality Weekly Report (MMWR) dates. As Pango variant designations were performed after completion of the study instead of in real time, and to prevent erroneous early detections of variants, variants were only assigned if the detection occurred after the date the variant was designated on the SARS-CoV-2 variants Github page (https://github.com/sars-cov-2-variants/lineage-proposals), otherwise the variants were grouped into earlier parent lineages. For clinical samples, weekly relative abundances were calculated using the number of samples that fell within a delineated parent lineage for each MMWR week. For air and wastewater samples, the de-mixed relative abundances for each parent variant were averaged across samples within each MMWR week, providing weekly mean relative abundances.

### Statistical comparison of air, wastewater and clinical data

Wastewater and air sample collection dates were grouped by MMWR week, with weeks defined as Sunday through Saturday. When multiple wastewater samples were collected from the same site within a given week, their values were averaged to produce a single wastewater concentration per site per week. Clinical percent positivity data for influenza A, influenza B, RSV, and SARS-CoV-2 were obtained from the Chicago Data Portal (https://data.cityofchicago.org). Percent positivity for air and wastewater samples was calculated based on PCR limit of detection (LoD) values. In air samples, the defined LoD was a Ct value of 38.3 for influenza A, 38.7 for influenza B, 37.9 for SARS-CoV-2, and 37.8 for RSV. In wastewater samples, the LoD was a viral load of 2025 copies/L for influenza A, SARS-CoV-2, and RSV and 2100 copies/L for influenza B. Wastewater viral load values were transformed to log-scale, with zero values changed to 100 for influenza A, influenza B, and RSV and 1,000 for SARS-CoV-2. Spearman correlation coefficients (α=0.05) were calculated and figures were generated using GraphPad Prism 10 for Windows (version 10.2.2). The absolute values of Spearman correlation coefficients were used in the heatmap, given the inverse relationship between Ct value and both wastewater concentration and clinical percent positivity. Comparison of air sample Cts across different site types was performed using a Kruskal-Wallis test with correction for multiple tests performed using Dunn’s multiple comparisons test using GraphPad Prism (version 10.2.3).

When comparing the SARS-CoV-2 sequencing genome coverage with Ct value or N1 gene copy concentration, the smoothed mean line was determined using the locally estimated scatterplot smoothing (LOESS) method with 95% confidence intervals. Two air samples were excluded as outliers because they had inexplicably low coverage despite low Ct values. Correlations between lineage relative abundances in clinical, air, and wastewater samples were determined using Spearman’s rank correlation coefficients.

To compare seasonal onset and peak across air, wastewater, and clinical data, “baseline” off-season periods were identified through visual inspection of clinical percent positivity data for influenza A, influenza B, RSV, and SARS-CoV-2. The mean and standard deviation was calculated for all weeks included in the off-season for all four pathogens in air, wastewater, and clinical data. An “above-baseline” threshold was defined as two standard deviations above the off-season mean. Seasonal onset was determined as the first week of two or more consecutive weeks exceeding the threshold within a given season. The seasonal peak was defined as the week with the highest observed value during that season.

Cross correlation functions (CCFs) were used to identify the lead or lag time (in weeks) between clinical percent positivity and environmental sample viral concentrations using Ct for air samples from all sites and air samples from EDs, and concentration from wastewater samples. Briefly, the *ccf* function in RStudio was used to evaluate the correlation strength between each environmental dataset with the clinical dataset synchronously and then offset by weekly positive and negative lag increments^18^. The weekly lag with the maximum CCF coefficient was reported as the lag between the datasets, with a negative value indicating the environmental trends preceded the clinical trends, and a positive value indicating the environmental trends lagged behind clinical trends. Data were analyzed across the entire study period as well as partitioned into independent years corresponding with respiratory virus seasons as follows: year 1= MMWR week 27 of 2023 through MMWR week 26 of 2024, year 2 = MMWR week 27 of 2024 through the end of the study period. MMWR week 49 was excluded from the wastewater correlations due to outlier wastewater concentrations that were several orders of magnitude above all other concentrations. Lag times were only reported if the CCF coefficient was statistically significant (p<0.05) by the *ccf* function.

### Data availability

SARS-CoV-2 sequence data are available in NCBI BioProjects PRJNA903175 (clinical data), PRJNA1337702 (air data), and PRJNA989260 (wastewater data).

## RESULTS

### Establishing and evaluating a citywide air surveillance program

Ten air samplers were positioned across the City of Chicago (Figure 1a). Five were placed in congregate living settings, three in hospital emergency departments (EDs), one in a hospital lounge area, and one in a health clinic. Healthcare and ED sites were targeted as community sites likely containing persons currently presenting with infectious diseases. Congregate living sites were selected based on history of respiratory virus outbreaks, vulnerable populations served, and ability to compare to other surveillance methods (e.g. clinical data and wastewater). Sites were selected across varying Chicago neighborhoods and Healthy Chicago Zones^27^ with a total of 776 weekly air samples were collected from all sites.

To validate the surveillance results of air monitoring program, air sampling data were compared to an existing wastewater surveillance program and local clinical case surveillance at both a citywide and single-facility scale (Figure 1b). These comparisons used 2,603 wastewater samples from eleven sub-sewershed sites (municipal sewers and pumping stations) located in Chicago, three treatment plants whose sewersheds include the city of Chicago, and 489 wastewater samples from a congregate facility that was also monitored by air surveillance. (Figure 1a). Chicago clinical case data was publicly available and represented as weekly aggregated laboratory tests voluntarily reported to CDPH by several hospital laboratories as well as two commercial laboratories that serve Chicago facilities as part of sentinel surveillance reporting^28^. A subset of Chicago clinical SARS-CoV-2 specimens were also subjected to whole genome sequencing as part of CDPH genomic surveillance activities, 4,916 from the study period were available for comparison^23^.

### Respiratory virus load and positivity correlate closely across citywide air, wastewater and clinical samples

Nucleic acids from respiratory viruses influenza A, influenza B, RSV, and SARS-CoV-2 were readily detected in air samples. 644 of 776 (83%) air samples were positive for at least one virus, with mean 1.7 viruses detected in each air sample. 358 of 412 (87%) of air samples from medical sites were positive for at least one virus compared to 286 of 364 (79%) air samples from congregate sites. Air sample Cts were lowest from EDs, with significantly lower mean Cts across all pathogens when compared to congregate sites (P<0.001) or across influenza A, RSV, and SARS-CoV-2 compared to other medical sites (P<0.0001) (Figure 2a). Congregate sites and non-ED medical sites had similar mean Cts across pathogens (P>0.05 for influenza A/B and RSV, P=0.16 for SARS-CoV-2) (Figure 2a).

**Figure 2:**
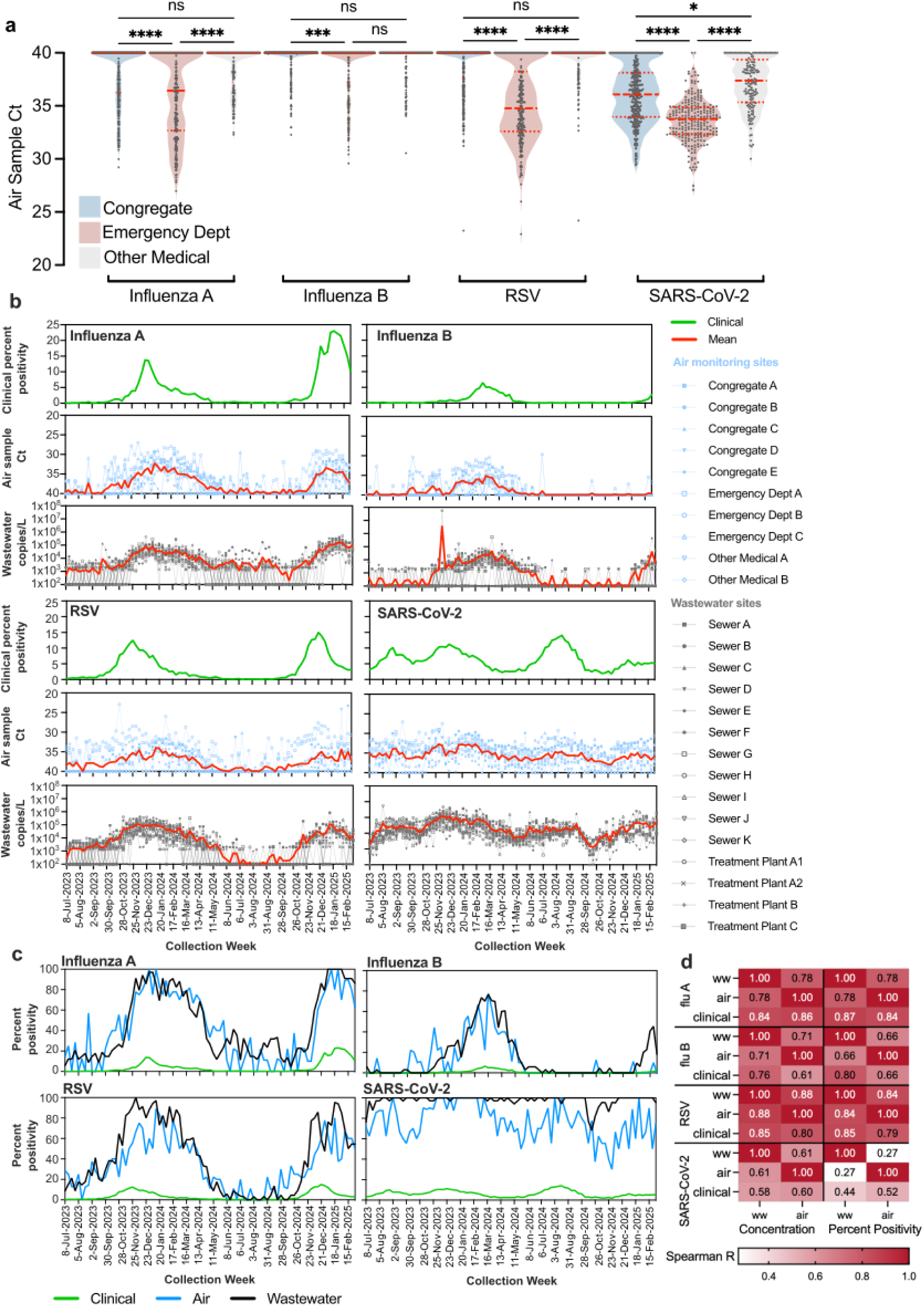
Respiratory virus positivity and load across citywide clinical, air, and wastewater samples. a) Air sample cycle threshold (Ct) values from samples collected from congregate (blue), emergency department (red) and other medical (grey) sites. Red dashed lines indicate median and dotted lines indicate quartiles. Significance of Kruskal-Wallis test shown above, adjusted for multiple comparisons: *, P<0.05; ***, P<0.001; *****, P<0.0001; ns, not significant. b) Clinical percent positivity (green), air sample Ct values (blue), and wastewater concentrations (grey) for influenza A, influenza B, RSV, and SARS-CoV-2. c) Percent positivity of clinical (green), air (blue), and wastewater (black) samples for influenza A, influenza B, RSV, and SARS-CoV-2. d) Heat map representing Spearman correlations between clinical, air, and wastewater samples using d/q PCR data (air sample Ct and wastewater concentration (copies/L)) and percent positivity. P<0.0001 for all comparisons.

Patterns of seasonal respiratory virus activity were clearly detected and overlapping across sample types (Figure 2b). Statistically significant correlations were observed between clinical, air, and wastewater samples when comparing both viral load (air and wastewater) and percent positivity (air, wastewater, and clinical) (Figure 2d). For example, influenza A detections began to rise in air, wastewater, and clinical specimens in October 2023, peaked in December 2023, and followed a similar pattern the following year, with an increase seen in November 2024 and a peak in January 2025 (Figure 2b). Influenza A clinical percent positivity showed a strong correlation with air sample Ct values (Spearman r: −0.86, p<0.0001) and wastewater concentrations (Spearman r: 0.84, P<0.0001). Influenza A viral loads in air and wastewater samples were also significantly correlated (Spearman r: −0.78, p<0.0001). Correspondingly, influenza A percent positivity was significantly correlated across all three surveillance methods (Spearman r: 0.78 - 0.87, p<0.0001, Figure 2c).

One advantage of air sampling is the ability to target sites relevant for specific pathogens. For example, children are particularly impacted by RSV; two air samplers in children’s hospitals, Emergency Dept A (open square) and Emergency Dept B (open circle, Figure 2b, Supplemental Figure 1), exhibited consistently higher RSV viral loads in air samples, and even showed sporadic RSV detections during periods of low clinical detection, showcasing the potential value of localized environmental surveillance in pediatric settings.

The weakest correlations among the three surveillance methods were observed in SARS-CoV-2 detection (Figure 2d; Spearman r:0.578, P<0.0001 for wastewater dPCR and r:0.603, P<0.0001 for air qPCR values compared to clinical percent positivity). Most wastewater sites were positive for SARS-CoV-2 during the study period, while there was more variability seen in air specimens. (Figure 2b-c). Overall, air surveillance at multiple aggregated sites well-reflected citywide wastewater and clinical case surveillance.

### Respiratory virus temporal trends via clinical, wastewater, and air surveillance

Environmental surveillance can serve as a leading indicator for pathogen trends. To compare the observed timing of respiratory virus trends across clinical, wastewater, and air data, we utilized maximum cross-correlation coefficients to identify the lead or lag time for environmental sample concentrations versus clinical percent positivity. We analyzed data across the entire study period as well as partitioned into two years corresponding with respiratory virus seasons (Table 3; Supplemental Figure 2). Lead/lag time varied across years within air and wastewater data but never exceeded a total difference of 3 weeks. Air samples from EDs were also analyzed independently of all air samples, as these showed the highest viral detections (Figure 2a).

**Table 3:** Cross correlation function lead/lag analysis.

| Pathogen | Dataset | CCF lag<br>(overall)<sup>a</sup> | CCF lag<br>(2023-2024) | CCF lag<br>(2024-2025) | CCF<br>value<br>(overall)<sup>b</sup> | CCF<br>value<br>(2023-2024) | CCF<br>value<br>(2024-2025) |
| --- | --- | --- | --- | --- | --- | --- | --- |
| Influenza A | Air (all)<sup>c</sup> | 0 | +2 | -1 | -0.73 | -0.82 | -0.97 |
|  | Air (ED)<sup>d</sup> | -1 | 0 | -1 | -0.72 | -0.76 | -0.94 |
|  | WW<sup>e</sup> | 0 | 0 | +1 | 0.92 | 0.91 | 0.93 |
| Influenza B | Air (all) | -1 | -2 | NA<sup>f</sup> | -0.82 | -0.82 | NA<sup>f</sup> |
|  | Air (ED) | 0 | -1 | NA<sup>f</sup> | -0.88 | -0.89 | NA<sup>f</sup> |
|  | WW | -1 | 0 | NA<sup>f</sup> | 0.77 | 0.84 | NA<sup>f</sup> |
| RSV | Air (all) | +4 | +4 | +2 | -0.71 | -0.71 | -0.76 |
|  | Air (ED) | 0 | 0 | 0 | -0.74 | -0.70 | -0.82 |
|  | WW | +3 | +2 | +3 | 0.82 | 0.92 | 0.82 |
| SARS-CoV-2 | Air (all) | +1 | +1 | +1 | -0.56 | -0.64 | -0.70 |
|  | Air (ED) | +1 | +1 | -1 | -0.61 | -0.62 | -0.67 |
|  | WW | 0 | +1 | NA<sup>g</sup> | 0.45 | 0.74 | NA<sup>g</sup> |

<sup>a</sup>Difference in weeks compared to clinical data, (-) lead, (+) lag
<sup>b</sup>Cross-correlation function value at indicated lead/lag week
<sup>c</sup>Data from all air samplers
<sup>d</sup>Data from air samplers placed in emergency departments
<sup>e</sup>Wastewater
<sup>f</sup>Data not reported because influenza B peak was not reached before the end of data collection
<sup>g</sup>Data not reported because no weeks reached a statistically significant positive CCF value.

For influenza A, air sample Cts from EDs led clinical percent positivity by 1 week across the entire study period; when divided into two years, EDs were aligned or led clinical data by one week for the 2023-2024 and 2024-2025 years, respectively. Air sample Cts from all sources and wastewater sample concentrations were aligned with influenza A clinical percent positivity across the entire study period, though showed some diversity when broken into years (air samples from all sites lagged by 2 weeks and led by 1 week, wastewater samples were aligned and lagged by one week for 2023-2024 and 2024-2025 years, respectively, Table 3, Supplemental Figure 2).

For influenza B, lags were only calculated across all sampling data and the 2023-2024 year, since the 2024-2025 peak had not been reached by the end of the study period. Across the study period, air samples from EDs were aligned with Influenza B clinical data and led clinical data by 1 week for the 2023-2024 year. Air samples from all sites across the study period led clinical data by 1 week and led clinical data by 2 weeks during the 2023-2024 year. Wastewater data overall led clinical percent positivity by 1 week and were aligned for the 2023-2024 year.

RSV samples showed the most disparate results between environmental sample types; air sample Cts from EDs were most closely aligned with clinical percent positivity, with no lag across the study period and in each year, while air sample Cts from all sites and wastewater sample concentrations both lagged behind clinical data by 4 and 3 weeks across the study, respectively (Table 3, Supplemental Figure 2). Finally, wastewater data was the most closely aligned with clinical SARS-CoV-2 percent positivity, and was aligned with clinical data across the study period and lagging by one week for the 2023-2024 year, although the 2024-2025 data did not reach statistically significant coefficients. Air sample Cts from all sites and from EDs both lagged behind clinical percent positivity for SARS-CoV-2 by 1 week across the study period. Yearly, air samples from all sites lagged by one weeks and air samples from EDs lagged and led by one week for 2023-2024 and 2024-2025 years, respectively, Table 3, Supplemental Figure 2).

We also compared timing of the onset and peak of each virus across both years. However, due to the long sampling period employed for air samples and grouping of wastewater and clinical data into corresponding week averages, we did not use smoothing analyses (e.g. rolling average) to reduce noise in the data, which is standard in similar wastewater studies^29^. Therefore, peak and onset lead and lag times were highly variable across air and wastewater compared to clinical data (range: leading by 5 weeks to lagging by 8 weeks, Supplemental Table 1). Generally, air samples from all sites and/or EDs led wastewater samples but lagged clinical samples, leading by a mean 1.2 weeks compared to wastewater and lagging by a mean 1.9 weeks compared to clinical samples for all comparisons.

### Whole-genome sequencing of SARS-CoV-2 from air and wastewater samples yielded high genome coverage

Whole-genome sequencing (WGS) of clinical and environmental samples provides an additional layer of surveillance – allowing for monitoring of viral variant trends over time as well as early alerts of new variants. We found that air samples are a good source of environmental SARS-CoV-2 genomes. 524 of 703 (74.5%) air samples surpassed our quality cutoff of 40% genome coverage. Air samples from ED sites most frequently reached our quality metric, with 223 of 235 (94.9%) reaching the cutoff. Air samples from other medical sites and congregate settings less frequently reached this cutoff, with 88 of 149 (59.1%) and 213 of 319 (66.8%) reaching the cutoff, respectively. Wastewater samples had higher rates of inclusion, with 1615 of 1700 (95.0%) surpassing the quality cutoff. This may be expected given the much larger populations included in wastewater specimens and likely higher SARS-CoV-2 gene copies in these samples. As expected, genome coverage increased with decreasing SARS-CoV-2 Ct values for air specimens and increasing SARS-CoV-2 concentrations for wastewater specimens (Supplemental Figure 3). Despite relatively small populations contributing to air samples versus wastewater samples (Table 1, Table 2), multiple SARS-CoV-2 lineages were detected in nearly all specimens, with a mean of 5.02 lineages (aggregated to most relevant variant families, see methods) detected in air samples. In comparison, mean 5.72 aggregated lineages were detected in wastewater samples.

### SARS-CoV-2 variant proportions were strongly correlated between different sample types

Both air and wastewater samples closely aligned with weekly SARS-CoV-2 variant proportions and trends in Chicago clinical samples (Figure 3a). Variant proportions were less variable across neighboring weeks in wastewater samples compared to air samples, with smaller populations surveilled by air samples possibly contributing to variability. Despite this, all 29 variants tracked in citywide clinical and wastewater samples were also detected in air samples from just 10 aggregated indoor locations (Figure 3a). Indeed, there were strong correlations between the relative abundances of the four most abundant aggregated variants in air and wastewater samples with clinical samples (Spearman r 0.68 - 0.89, p<0.0001 for air samples and 0.85 - 0.92, p<0.0001 for wastewater samples, Figure 3b). Relative abundances of these variants were also strongly correlated between air and wastewater samples (Spearman r 0.70 - 0.80, p<0.001). Other less abundant variants (maximum relative proportion not exceeding 45% in clinical data) showed varying correlations from very strong to no correlation (range −0.29-0.91, Supplemental Figure 4). Air and wastewater variant abundance correlation strength with clinical samples increased with increasing maximum variant abundance in clinical samples (Spearman r=0.56, p<0.01, and r=0.72, p<0.0001 respectively, Supplemental Figure 4). Wastewater correlations with clinical samples were higher than air in 27 of 29 variants (Supplemental Figure 4).

**Figure 3:**
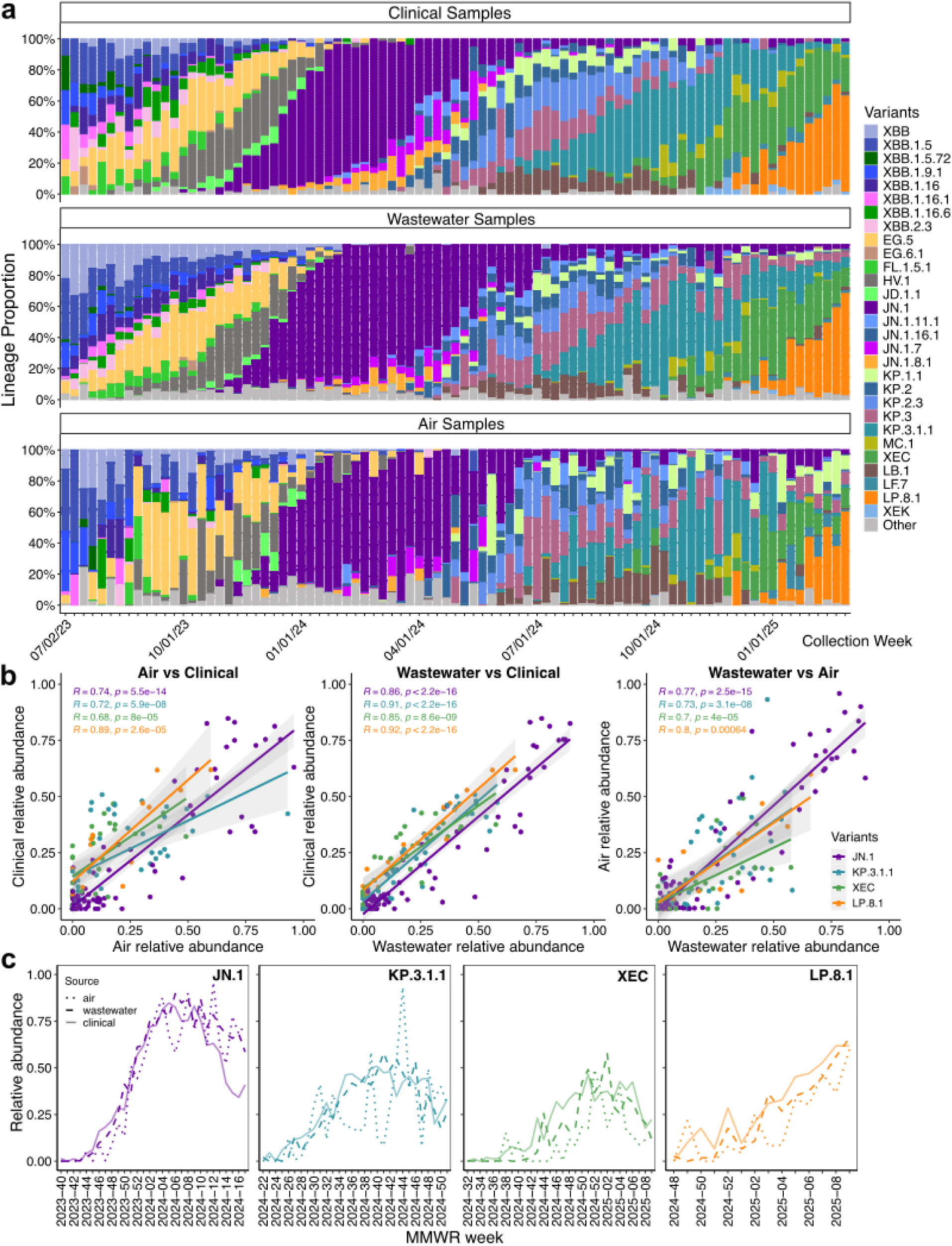
SARS-CoV-2 lineages detected in citywide clinical, air, and wastewater samples. a) Stacked bar charts representing the weekly relative abundance of SARS-CoV-2 variants in clinical, wastewater, and air samples. Variants were grouped into parent lineages as described in the Methods. Mean relative abundances per week are shown for wastewater and air samples. b) Correlation plots of the relative abundance of four common variants in weekly air vs clinical samples, wastewater vs clinical samples, and wastewater vs air samples. Linear regression line for each variant and 95% confidence interval (shaded) shown. Strength and significance of correlation was determined by Spearman correlations. c) Emergence of the four most common variants in air (dotted lines), wastewater (dashed lines), and clinical (solid lines) samples. Up to the first 30 weeks after first detection and lineage designation (whichever came last) shown.

### Monitoring SARS-CoV-2 variant emergence in air and wastewater

Of the 29 SARS-CoV-2 variants tracked here, 18 of 29 emerged during this study, meaning they were not observed in any sample type until after the first three weeks of the study (Supplemental Table 2). Some of these variants were first detected in one sample type only: 3/18 variants (MC.1, LF.7, and XEK) were first detected only in air samples, 8/18 (JD.1.1, KP.2, KP.1.1, JN.1.7, JN.1.11.1, KP.3.1.1, XEC, and LP.8.1) were first detected in only wastewater samples, and 2/18 (JN.1 and JN.1.8.1) were first detected in only clinical samples (Supplemental Table 2). The remaining variants were first detected in multiple sample types simultaneously (2/18 air and wastewater, 1/18 wastewater and clinical, 2/18 all three).

The tracked variants emerged at similar times in all sample types – clinical, air, and wastewater (Figure 3c). Wastewater most reliably detected variants early; when variants were first detected in another sample type, the variant was detected in wastewater a median of 1 week later (range 1-5 weeks, Supplemental Table 2, Supplemental Figure 5). Air and clinical detections were more irregular; when variants were first detected in other sample types, variants were detected in air and clinical samples a median of 1 and 4 weeks later (ranges of 1-14 and 1-20 weeks, respectively).

### Efficient respiratory virus surveillance of a single institution using indoor air sampling

Localized respiratory virus surveillance may be useful for outbreak monitoring or surveillance of high interest sites, e.g. points of entry for infectious diseases or sites where infectious diseases must be quickly controlled. One large congregate living institution with multiple buildings was sampled by both air and wastewater programs during this study. This site contained two air sampling sites in separate buildings and eight wastewater sampling sites targeting separate buildings. Both buildings containing an air sampling device were also monitored by wastewater sampling, although populations contributing to the environmental sample types may have differed. Air sampling devices were placed in rooms that were a gathering point for individuals who were transient in the room and weren’t necessarily housed in that building and who may or may not have used the bathroom there. Wastewater samples may have more closely reflected individuals housed in the building.

Although facility-level clinical data were not available, patterns of respiratory virus positivity seen at the citywide level were seen in both air and wastewater from this single institution for seasonal respiratory viruses influenza A, influenza B, and RSV (Figure 4a). Like at the citywide scale, fluctuations in SARS-CoV-2 levels were less discernable compared to clinical trends (Figure 4a). Air samplers at this facility efficiently reproduced citywide clinical trends, with each air sampler showing overlapping periods of viral detections compared to citywide clinical data. Facility air samplers showed detectible influenza A during a period of increased citywide clinical activity from 2023-11-25 to 2024-05-11, with 46 of 47 (98%) air samples showing detectable influenza A during this time (Figure 4a). Facility wastewater detections were less sensitive, with only 56 of 154 (36%) of wastewater samples showing influenza A detections during this time. Similarly, during a period of increased citywide influenza B clinical positivity, although relatively low during 2023-2024 season, influenza B was found in 30 of 31 (97%) facility air samples collected between 2024-01-20 to 2024-05-11 compared to 15 of 91 (16%) facility wastewater samples (Figure 4a). Both facility air samplers were well-correlated with Chicago clinical percent positivity for influenza A, influenza B, and RSV (mean Spearman r=0.74 and 0.67 for Site-A and Site-B, respectively, range 0.67-0.79 and 0.60-0.75), while facility wastewater sites were less well-correlated (mean Spearman r=0.30, range −0.05 to 0.66. Figure 4b).

**Figure 4:**
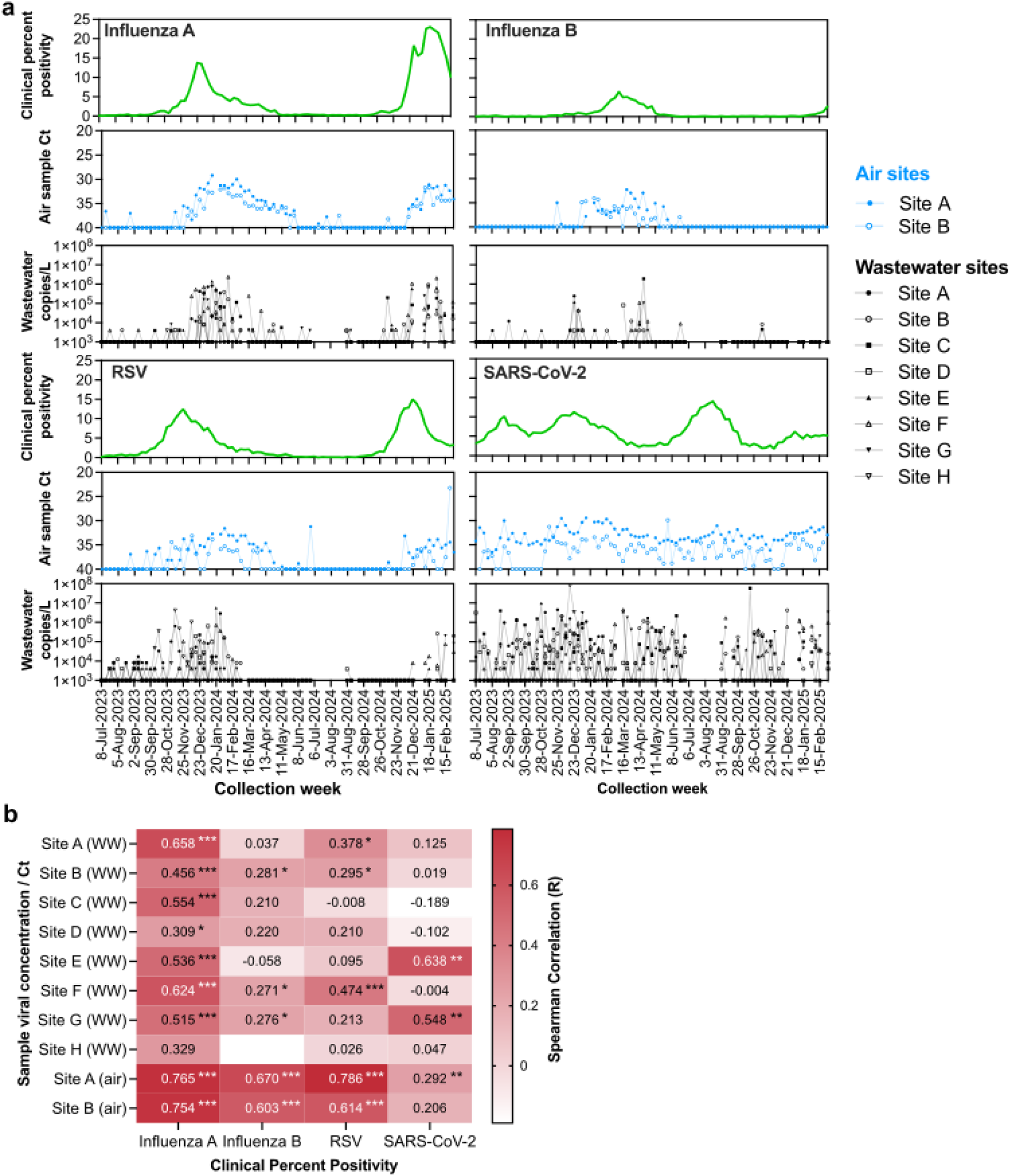
Monitoring respiratory virus activity in a single institution. a) Air sample Ct values (blue) and wastewater concentrations (black) for respiratory viruses influenza A, influenza B, RSV, and SARS-CoV-2 are compared to clinical percent positivity (green). Different sampling sites are denoted with differing node shapes. Undetectable values are plotted at 40 for air samples and 1000 for wastewater samples. b) Heatmap of Spearman R values for air and wastewater sites (rows) compared to clinical percent positivity for respiratory viruses (columns). Asterisks indicate significance values: * P<0.05; ** P<0.01; *** P<0.0001. Spearman R was unavailable for Site H as it had no positive detections for RSV.

### Environmental samples from a single site mirror citywide clinical SARS-CoV-2 variant trends and may detect local variations

We conducted SARS-CoV-2 WGS on samples from both air surveillance sites (Air Site A and Air Site B) and Wastewater Site B from this congregate living institution to observe relative abundances of SARS-CoV-2 variants at the institution level (Figure 5). We obtained data above our genome coverage cutoff for 79/87 weeks from Air Site A, 48/87 weeks from Air Site B, and 38/87 weeks for Wastewater Site B. Despite the small number of sampling sites and data surpassing inclusion cutoffs, we observed generally similar trends between the air and wastewater samples and citywide clinical samples, particularly in more prevalent variants (Supplemental Figure 5). Relative abundance of JN.1 in air and wastewater samples was moderately correlated with the variant’s relative abundance in citywide clinical samples (Spearman r of 0.49, p<0.05 and 0.57, p<0.01, respectively) but were strongly correlated with one another (Spearman r of 0.76, p<0.0001).

**Figure 5.**
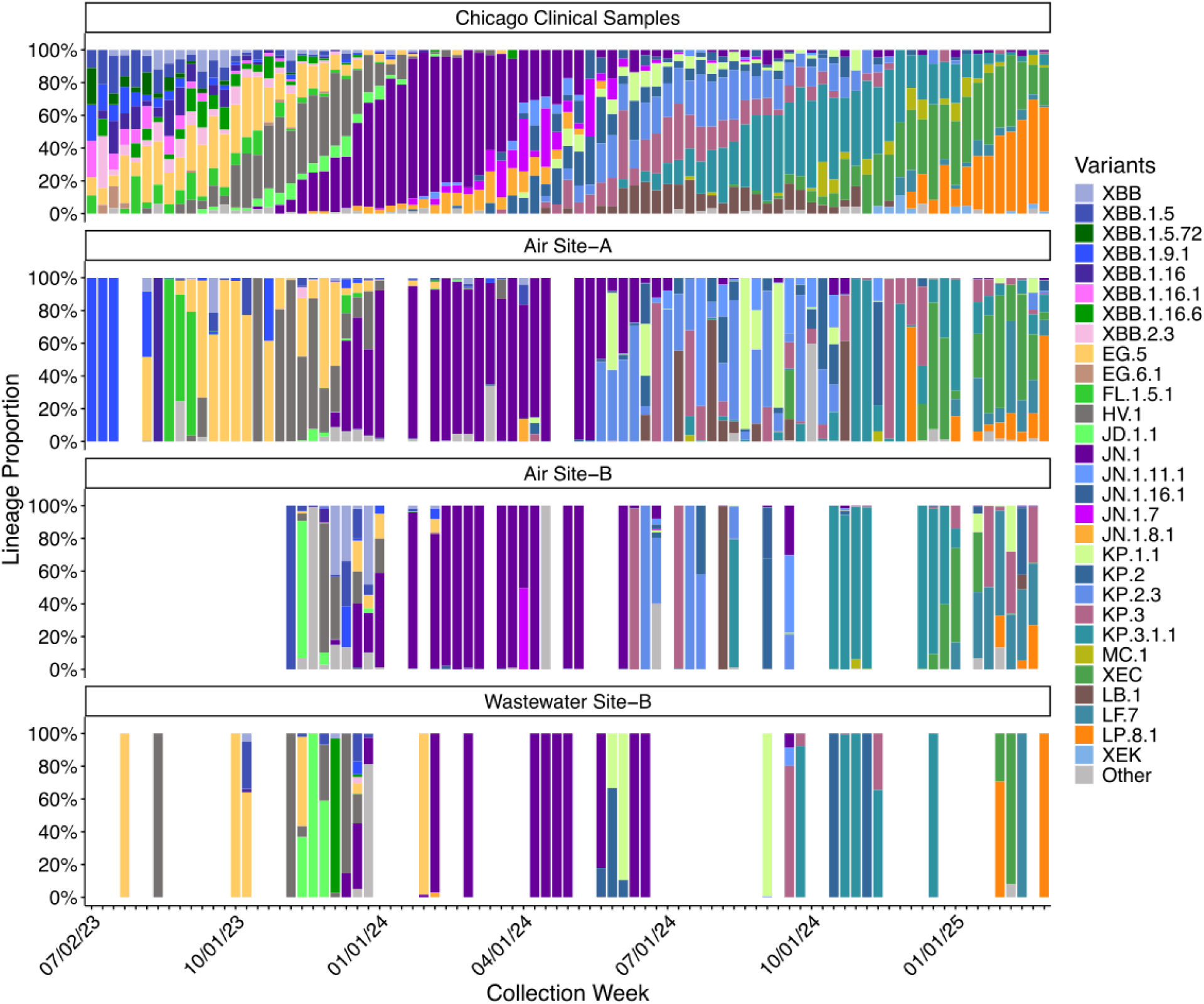
Monitoring SARS-CoV-2 lineages in a single institution. Stacked bar charts representing the weekly relative abundance of SARS-CoV-2 variants in clinical samples from Chicago and air and wastewater samples from a single institution. Variants were grouped into parent lineages as described in the Methods.

We also observed divergence from citywide clinical trends in air samples from this institution. For example, in 2023 MMWR weeks 46 (Air Site B) and 46-48 (Wastewater Site B), we observed a high proportion of JD.1.1 that wasn’t seen in citywide clinical cases (Figure 5). These two sites sampled the same building, suggesting we may have observed localized transmission of JD.1.1, though it is unknown if this was due to increased cases at this site or one/few infected persons with high shedding rates^30^. We observed similar peaks in 2024 MMWR weeks 22-25 and 34-37, where elevated levels of KP.1.1 were observed in Air Site A and Wastewater Site B, but not citywide clinical samples or Air Site B. Again, while our clinical sampling cannot confirm whether these incidences were local increases of these lineages, in both cases the variant surges were detected in both air and wastewater samples. Overall, these data highlight how air sampling may be well-suited for building-scale environmental surveillance (Table 4).

**Table 4:**
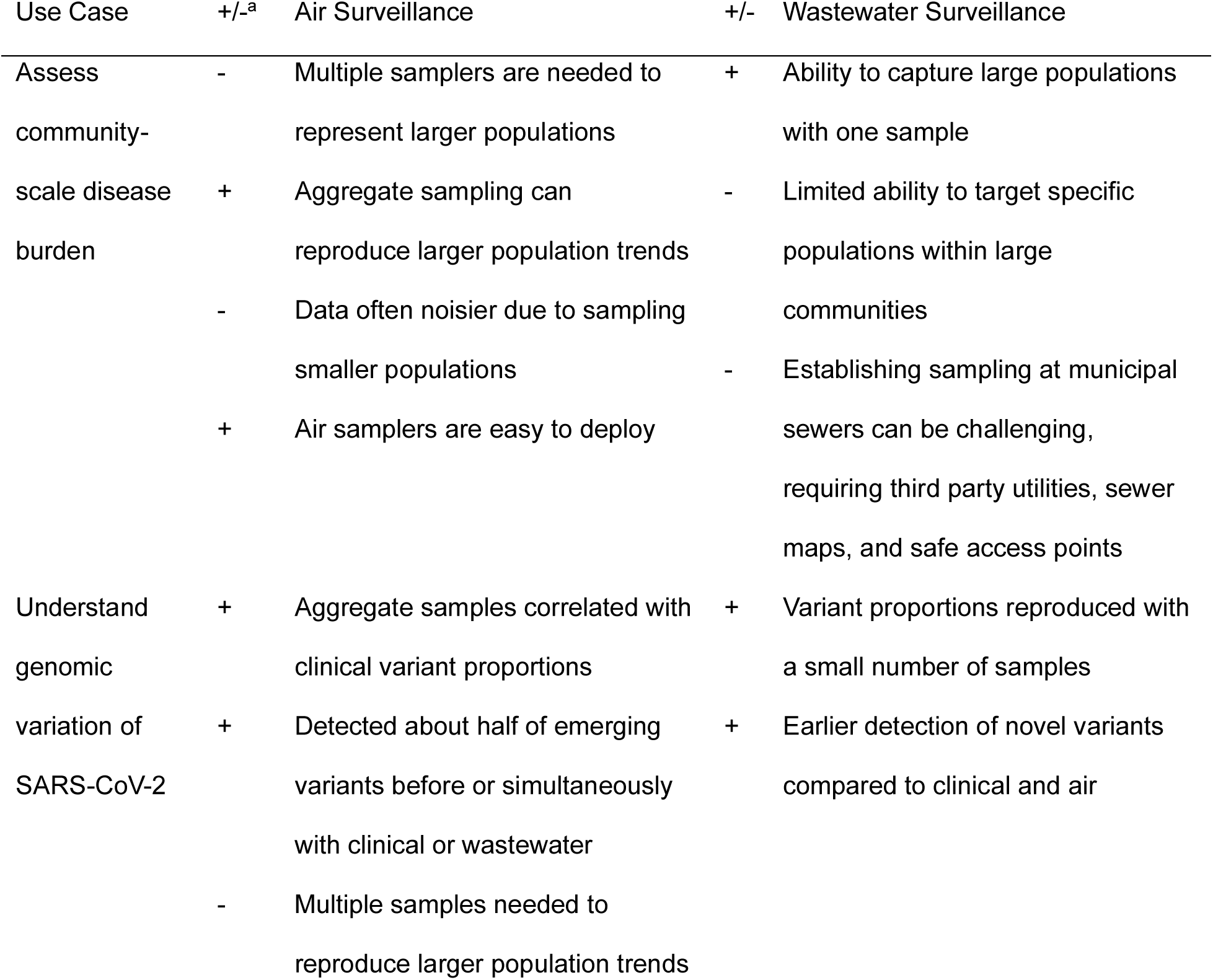
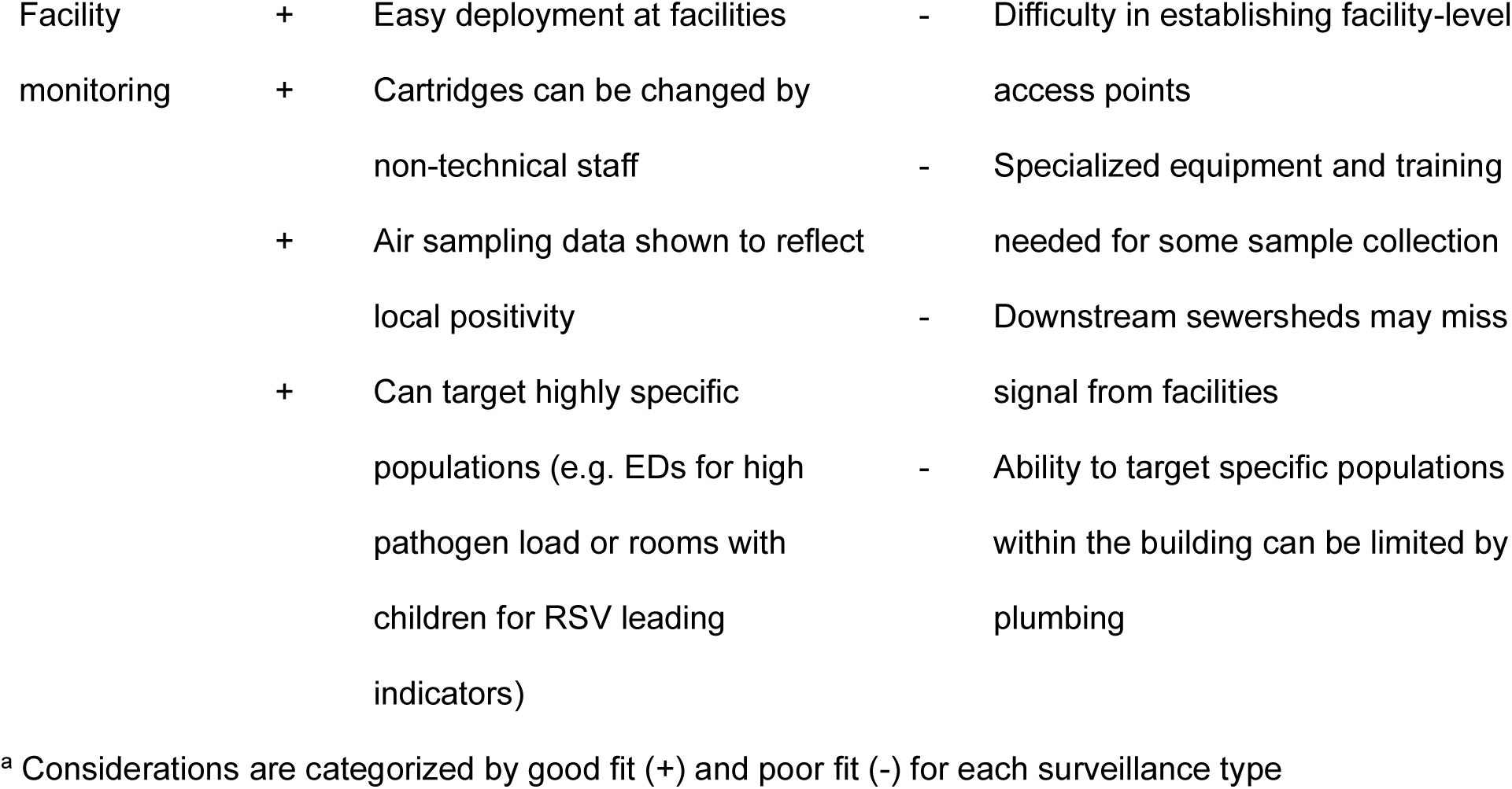
Environmental Surveillance Use Cases: Air and Wastewater Perspectives.

## DISCUSSION

Many studies have shown that respiratory pathogens can be detected in air samples from indoor settings, but how well these detections correlate to other established surveillance methods such as wastewater or local clinical cases is less well known^12–14,31–33^. Further, combining data from multiple indoor air sampling sites for community-scale pathogen surveillance has not been validated. Here, we placed air samplers at several indoor locations around a large city to establish local surveillance and determine whether air sampling could accurately represent clinical case trends and serve as an additive tool for local environmental surveillance. To be useful for public health surveillance programs, air surveillance must be representative of cases in the sampled location and have additive value in jurisdictions already employing traditional or wastewater surveillance methods. Here, we found that air samplers at just 10 medical and congregate living locations across the city of Chicago produce aggregate viral activity data that is well-correlated to clinical case and wastewater data for all respiratory viruses analyzed. Additionally, these air samples yielded SARS-CoV-2 lineage proportions that are well-correlated to clinical case and wastewater data, suggesting that air sampling may have applications for infectious disease surveillance as well as genomic surveillance.

We targeted two types of facilities for their distinct surveillance use cases: i) medical institutions, to capture individuals with infectious diseases and ii) congregate living facilities, which have historically faced pathogen outbreaks. We found that air samples from EDs showed the highest respiratory viral loads. Air samples from non-ED medical institution sites (a specialty clinic and an acute care hospital restaurant/lounge) had similar viral loads in air samples compared to congregate settings (group homes and a long term care facility), suggesting that EDs might be ideal targets for sensitive detection of respiratory pathogens and other community sites may serve as well as non-ED medical sites. We found that air surveillance results from a single congregate living institution aligns with citywide clinical percent positivity, suggesting that access to medical institutions may not be needed to perform air surveillance that is representative of local cases. However, this congregate living facility contained a large population with rapid turnover; facilities with consistent or isolated populations may be less representative of local cases. Others testing multiple community sites found highest respiratory virus positivity in air samples from nurseries, schools, and other sites populated with young children or adolescents, suggesting that these may be other ideal candidates for air surveillance programs^12,13^.

There are a few gaps in existing environmental surveillance programs that air sampling may fill. A key advantage of air surveillance is the ease of sampler deployment and sample acquisition. Air samplers can be placed within minutes, and cartridge exchanges can be done by non-technical personnel. In contrast, wastewater collection, especially at private institutions, requires coordination of individuals with specialized training and equipment^34^. Additionally, access to municipal sewers can be challenging, requiring coordination with third party utilities, availability of sewer maps, and identification of appropriate and safe access points. Building sampling can require confirmation of effluent at the access point via dye tracer tests, coordination with building engineers, and can be contaminated by organisms in biofilms for bacterial pathogens, which may not be representative of organisms in building inhabitants^35^. For these reasons, establishing a wastewater sampling site can often take months^35^. Air samplers may therefore be useful for rapid deployment in outbreak responses of targeted populations.

Though wastewater data is likely still optimal for large population-level surveillance, air surveillance provides many enhanced benefits when targeting sub-populations. We found air sampling in a single facility was more sensitive and well-correlated with community percent positivity than wastewater for influenza A, influenza B, and RSV, suggesting that air surveillance may be preferred over wastewater when targeting small populations for respiratory viruses. Healthcare facilities could use unit-specific data to inform changes to PPE requirements or other infection prevention practices during periods of increased pathogen transmission. Air sampling could also be used to enhance coverage in sub-populations with historically low-health care seeking behaviors and/or health care access, or those that do not contribute to accessible wastewater, e.g. diapered individuals or in rural areas that are unsewered. Congregations with low vaccination rates could be prioritized for surveillance of vaccine-preventable diseases during outbreaks, and if disease trends in these locations vary from population trends, targeted mobile testing or vaccination efforts could be deployed.

Here, we observed that a small number of samplers can still successfully reflect clinical trends, with aggregated data from just 10 air samplers representing ∼1000-fold smaller populations than wastewater reproducing respiratory virus trends and SARS-CoV-2 variant proportions. The ability to split up community-scale air data by individual sampling locations might provide extra information that is limited in surveillance of large populations like wastewater treatment plants. This could give insight into which sub-populations are being differentially affected by pathogens (e.g. children’s hospitals versus adult hospitals versus young adults in college clinics) or identify the precise neighborhood source of an outbreak without additional sampling or source tracing.

Air versus wastewater surveillance may be selected based on pathogen. Individual respiratory, gastrointestinal, and skin-associated pathogens are differentially shed into the air and wastewater and this could impact ideal surveillance strategy. Air surveillance may not be as effective for mild but symptomatic infections that drive infected individuals to recover at home, but may effectively capture very mild or asymptomatic infections that do not limit activity or more severe infections that bring infected people to EDs where samplers may capture them.

One of the highlights of wastewater surveillance has been its use as a leading indicator for clinical case trends. Here a cross-correlation lead/lag analysis showed varying temporal trends, with air, clinical and/or wastewater data leading for individual pathogens and seasons. Environmental data are thought to lead clinical data due to virus shedding prior to symptom onset. If early warning is desired, air surveillance programs could target specific populations that are more likely to serve as leading indicators. For example, two of three EDs subjected to air sampling in this study were in children’s hospitals. We found that air RSV data from EDs coincided with clinical data, while air data from all sites and wastewater data lagged by 2 or more weeks. RSV detections in non-ED air and wastewater data may be limited by a lack of children in the sampled sites for air and diapered individuals for wastewater. Air sampling in non-medical buildings with children may further precede ED RSV detection if capturing pre-symptomatic individuals. Only sampling at medical sites with symptomatic patients may diminish the early detection potential of air samples. For influenza B, air data from emergency departments lagged 1 week behind air data from all sampling sites, perhaps because pre-symptomatic individuals were sampled in non-medical settings.

In addition to clinical trends, wastewater also serves as a leading indicator for novel SARS-CoV-2 variants^10^. Here, novel SARS-CoV-2 lineages were most frequently detected first in wastewater data, likely due to the large populations contributing to these samples. However, despite a much smaller population surveilled, nearly half of newly emerging variants were detected first or simultaneously in air data compared to wastewater, perhaps because the newly emerging variants become too dilute in wastewater sites serving a large population. Importantly, while “leading indicator” analyses are performed on retrospective data, the cadence of data reporting should be factored into the utility of the early indication for public health purposes. Clinical case diagnostic data is often rapidly tested and reported and actual lead times of environmental samples may be decreased by weekly or biweekly testing. Reporting lag maybe be more similar for results that include WGS like SARS-CoV-2 variant detection. Here, air samples were exchanged weekly for cost efficiency but greatly reduced their utility as an early indicator.

This study had some limitations. First, air sampling locations were not necessarily representative of the local population. Air sampling locations were selected in part based on placement feasibility, such as sites with existing partnerships with the health department or available resources for air sample collection. While samplers were able to be placed throughout the city, not all regions within Chicago were able to be included in this timeframe. Given the promising proof-of-concept data provided by this study, an air surveillance program containing locations that are more representative of the population could be developed. Room occupancy data, e.g. how many individuals were present and where they reside, could be very useful when assessing the representativeness of the sampling locations, but were not able to be collected as part of these studies. Air quality metrics like indoor CO_2_ concentration and whether the room contained air filtration systems, which are risk factors for pathogen load in air samples, were not collected as part of this study and were not considered when selecting air sampling locations^13^. Similarly, air sampling locations within buildings were chosen based on convenience and accessibility for staff, e.g. electrical outlet access and placement away from busy areas, rather than coordinating with building air handling for optimal sampling locations.

Overall, air sampling can provide environmental surveillance data that well-represents clinical and wastewater data at the building and community scale. It is an additive surveillance method that can be quickly deployed to targeted populations and sites and operated by non-technical individuals. Clinical laboratory testing has declined since the expiration of COVID-19 public health emergency. Environmental surveillance is needed to efficiently surveille populations where clinical surveillance is not feasible or available. Further, environmental sampling and sequencing may be ideal for detecting emerging pathogens. As such, air surveillance could become an essential tool for public health surveillance, complementing wastewater surveillance as a finer-scale and quicker-to-deploy targeted surveillance method. Because the acceptance of novel surveillance data can be slow by the public and/or public health institutions, additional opportunities for air surveillance studies are needed to prove its utility and gain confidence in the method.

## Data Availability

Sequence data are available in NCBI BioProjects PRJNA903175 (clinical data), PRJNA1337702 (air data). PRJNA989260 (wastewater data).

## Acknowledgements

We thank our colleagues at the Chicago Department of Public Health for their invaluable support and review of this work, specifically Stephanie R. Black, MD, MSc; Peter DeJonge, PhD Michelle Funk, DVM, MPH; Colin Korban, MPH; Peter Ruestow, PhD, and Haifa Wahbeh, MBA, CFFE.

We thank the Genomics and Microbiome Core Facility at Rush University Medical Center for laboratory processing of air samples, qPCR analysis and SARS-CoV-2 library preparation and sequencing, especially Kevin Kunstman, Giancarlo Balangue, Felix Araujo-Perez, Jeremy Kahsen, and Marisol Dominguez.

We thank Drs. David and Shelby O’Connor at the University of Wisconsin Madison for sharing air sampling expertise in support of this study.

We acknowledge the Illinois Department of Public Health, Metropolitan Water Reclamation District of Greater Chicago and Chicago Department of Water Management for their contributions and support of wastewater surveillance in Chicago and Cook County.

Finally, we gratefully thank air sampling sites for weekly cartridge exchanges, and support of the program, specifically Larry K. Kociolek, MD and Megan E. Reyna, BA at the Division of Pediatric Infectious Diseases, Ann & Robert H. Lurie Children’s Hospital of Chicago; Nidhi S. Undevia, M.D. and Lisa A. Duffner, BA, BS at RML Specialty Hospital Office of Clinical Research, Hinsdale, IL; Priscilla Ware, MD at Cermak Health Services and Juvenile Temporary Detention Center; Annie Chambers, B.A. at the Cook County Sheriff’s Office; Chad Zawitz, MD at Cook County Health; Allison H. Bartlett, MD, MS and David Zhang, MD at Comer Children’s Hospital, University of Chicago Medicine; Michael Gottlieb, MD, at the Department of Emergency Medicine, Rush University Medical Center; and the staff at the Chicago Department of Public Health Lakeview Sexual Health Clinic.

## Author contributions

Conceptualization: H.J.B, V.E.M, A.K.; Methodology: H.J.B, E.P.N, S.B., R.P, S.G., S.M.O, S.J.G, V.E.M,; Formal analysis: H.J.B, E.P.N, S.B.; Investigation: H.J.B, E.P.N, S.B., R.P, S.G., S.M.O., D.S.G., C.L., A.H., M.L.J., R.M.; Resources: R.J.B, S.S., D.F., A.K.; Writing – Original Draft: H.J.B, E.P.N, S.B. V.E.M, A.K.; Visualization: H.J.B, E.P.N, S.B.. A.T.; Supervision: H.J.B, R.P, S.M.O, S.J.G, M.H., S.R.B., A.K.; Funding acquisition: S.R.B., A.K.

