## Supplementary Information for "Citywide indoor air sampling mirrors wastewater and clinical case surveillance of respiratory viruses"

### Supplementary Figures

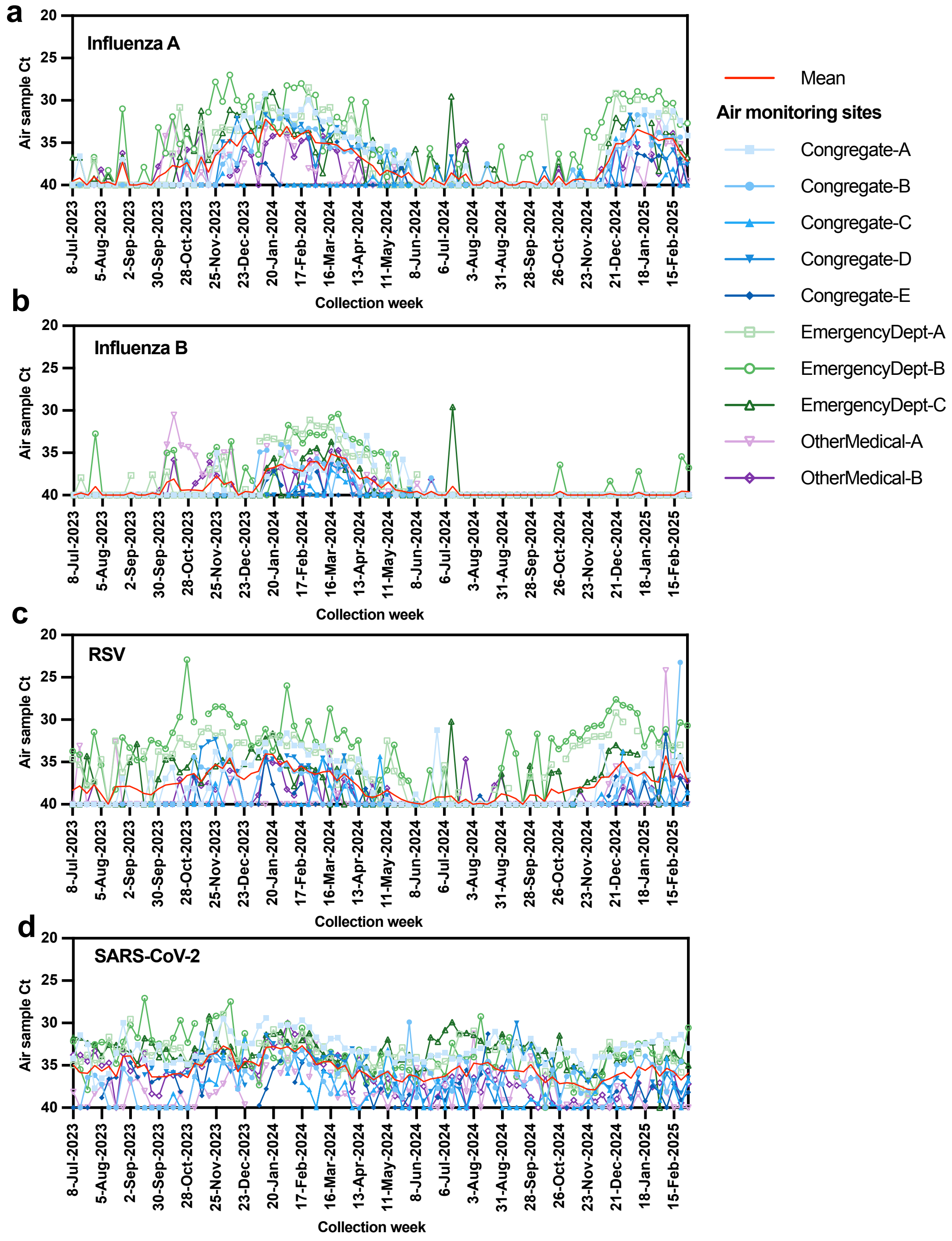


**Supplemental Figure 1.** Air sample Ct values (as in Figure 2B-E) colored by congregate (blue), emergency department (green) and other medical (purple) sampling sites for a) influenza A, b) influenza B, c) RSV, and d) SARS-CoV-2.

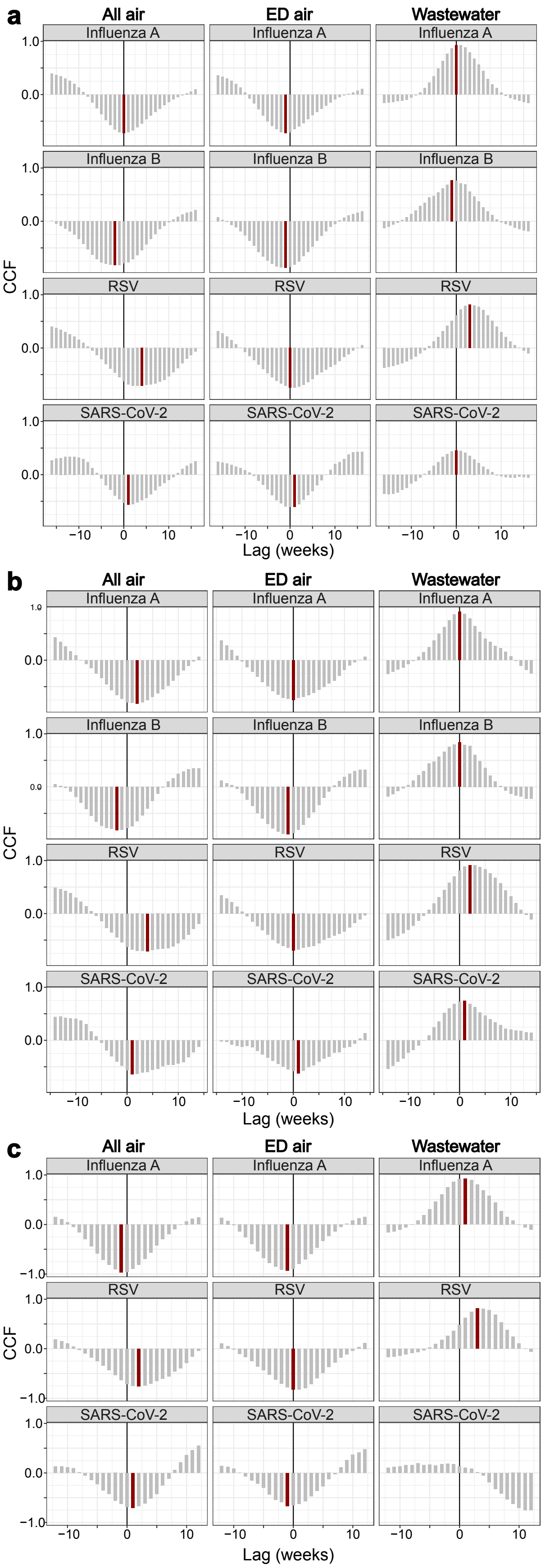

a
All air
ED air
Wastewater
Influenza A
Influenza B
RSV
SARS-CoV-2
CCF
1.0
0.0
−10
0
10
Lag (weeks)
b
All air
ED air
Wastewater
Influenza A
Influenza B
RSV
SARS-CoV-2
CCF
Lag (weeks)
c
All air
ED air
Wastewater
Influenza A
RSV
SARS-CoV-2
CCF
−1.0
Lag (weeks)

**Supplemental Figure 2:** Cross-correlation function (CCF) coefficients across a) the entire study period, b) year 1 (MMWR week 27 of 2023 through MMWR week 26 of 2024), or c) year 2 (MMWR week 27 of 2024 through the end of the study period). The dark red bar indicates the week lag with the maximum CCF coefficient; this coefficient is only shown if the CCF coefficient was statistically significant ($p<0.05$) by the *ccf* function.

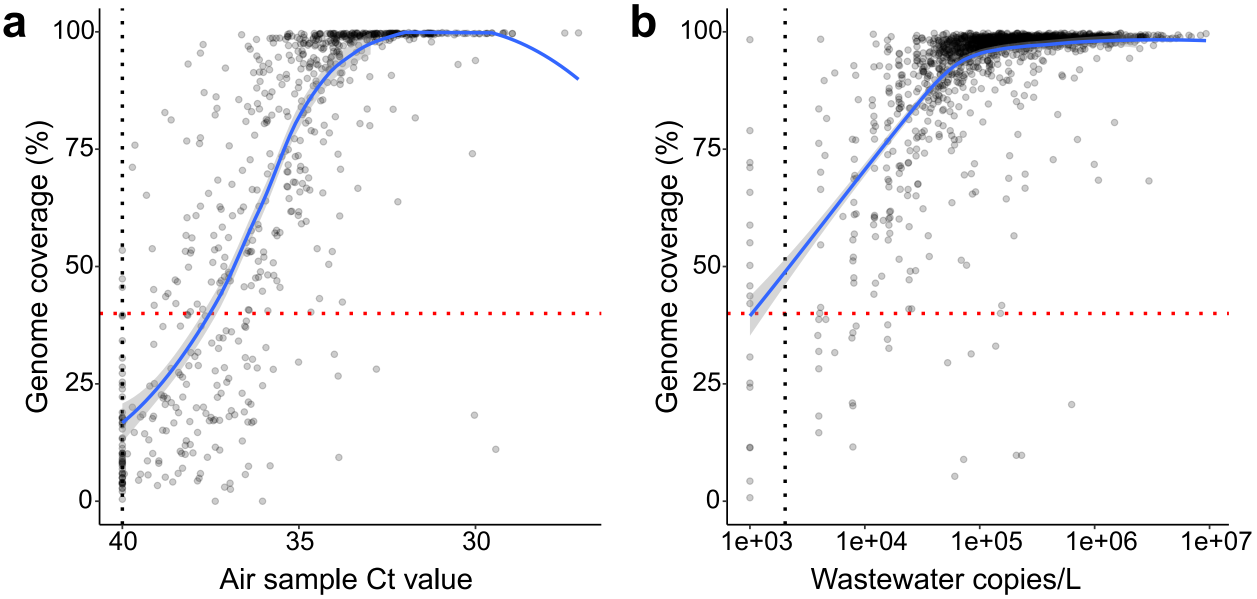


**Supplemental Figure 3.** SARS-CoV-2 genome coverage from the Freyja pipeline versus the a) Ct value for air samples or b) SARS-CoV-2 N1 gene copies/L from dPCR. The blue line is a smoothed trend line using the locally estimated scatterplot smoothing (LOESS) method with 95% confidence intervals in grey. The two points in the bottom right hand corner of a) were excluded as outliers from trend line calculations. The vertical black dotted line is the limit of detection (LOD) for each sample type, and the horizontal red dotted line is the 40% genome coverage cutoff used for inclusions in further analysis.

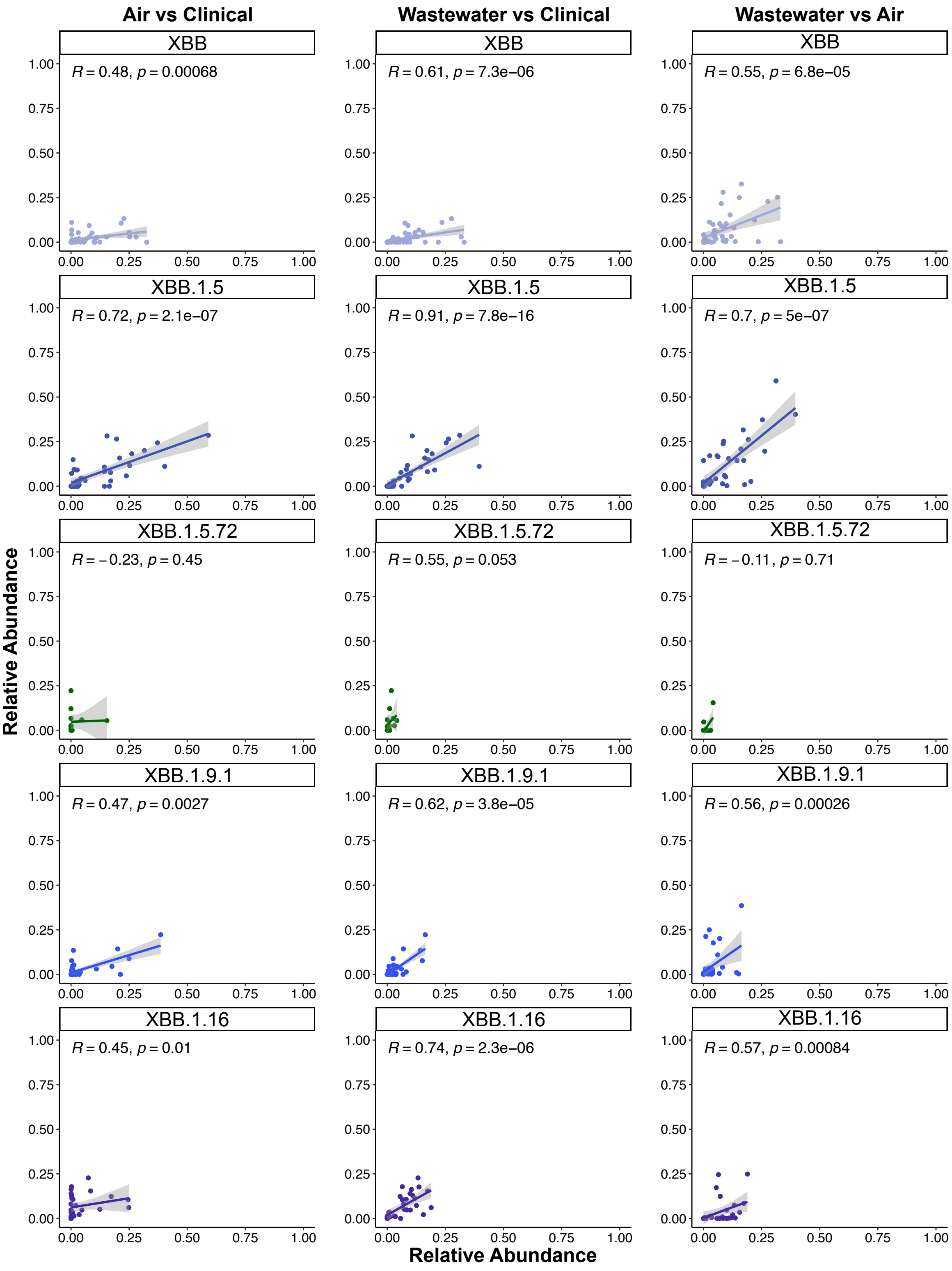

Air vs Clinical
Wastewater vs Clinical
Wastewater vs Air
XBB
R = 0.48, p = 0.00068
R = 0.61, p = 7.3e−06
R = 0.55, p = 6.8e−05
XBB.1.5
R = 0.72, p = 2.1e−07
R = 0.91, p = 7.8e−16
R = 0.7, p = 5e−07
XBB.1.5.72
R = −0.23, p = 0.45
R = 0.55, p = 0.053
R = −0.11, p = 0.71
XBB.1.9.1
R = 0.47, p = 0.0027
R = 0.62, p = 3.8e−05
R = 0.56, p = 0.00026
XBB.1.16
R = 0.45, p = 0.01
R = 0.74, p = 2.3e−06
R = 0.57, p = 0.00084
0.00
0.25
0.50
0.75
1.00
Relative Abundance
Relative Abundance

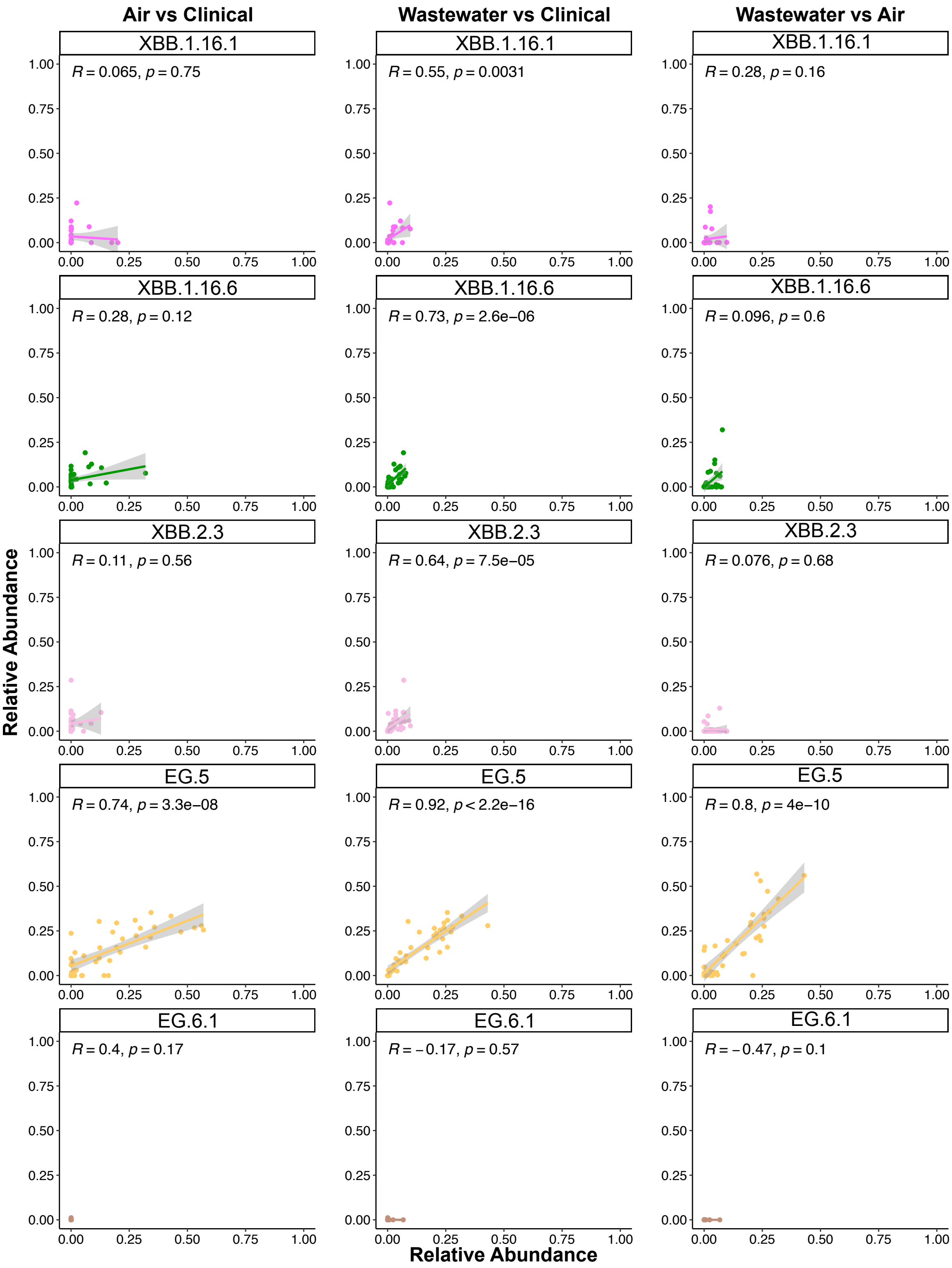
Air vs Clinical
Wastewater vs Clinical
Wastewater vs Air
XBB.1.16.1
R = 0.065, p = 0.75
R = 0.55, p = 0.0031
R = 0.28, p = 0.16
XBB.1.16.6
R = 0.28, p = 0.12
R = 0.73, p = 2.6e−06
R = 0.096, p = 0.6
XBB.2.3
R = 0.11, p = 0.56
R = 0.64, p = 7.5e−05
R = 0.076, p = 0.68
EG.5
R = 0.74, p = 3.3e−08
R = 0.92, p < 2.2e−16
R = 0.8, p = 4e−10
EG.6.1
R = 0.4, p = 0.17
R = −0.17, p = 0.57
R = −0.47, p = 0.1
Relative Abundance
Relative Abundance
0.00
0.25
0.50
0.75
1.00

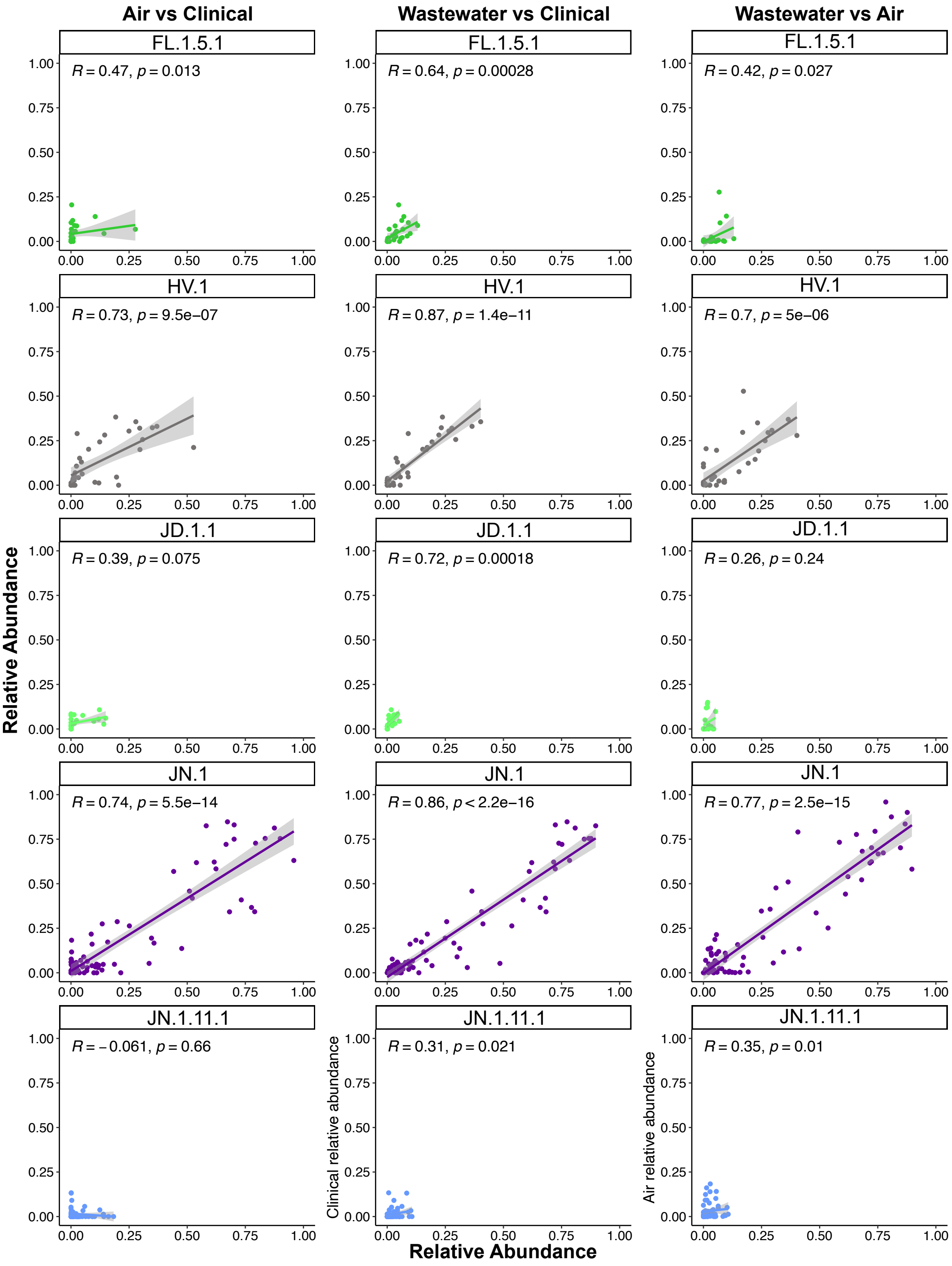

Air vs Clinical
Wastewater vs Clinical
Wastewater vs Air
FL.1.5.1
R = 0.47, p = 0.013
R = 0.64, p = 0.00028
R = 0.42, p = 0.027
HV.1
R = 0.73, p = 9.5e−07
R = 0.87, p = 1.4e−11
R = 0.7, p = 5e−06
JD.1.1
R = 0.39, p = 0.075
R = 0.72, p = 0.00018
R = 0.26, p = 0.24
JN.1
R = 0.74, p = 5.5e−14
R = 0.86, p < 2.2e−16
R = 0.77, p = 2.5e−15
JN.1.11.1
R = −0.061, p = 0.66
R = 0.31, p = 0.021
R = 0.35, p = 0.01
Relative Abundance
Clinical relative abundance
Air relative abundance
Relative Abundance

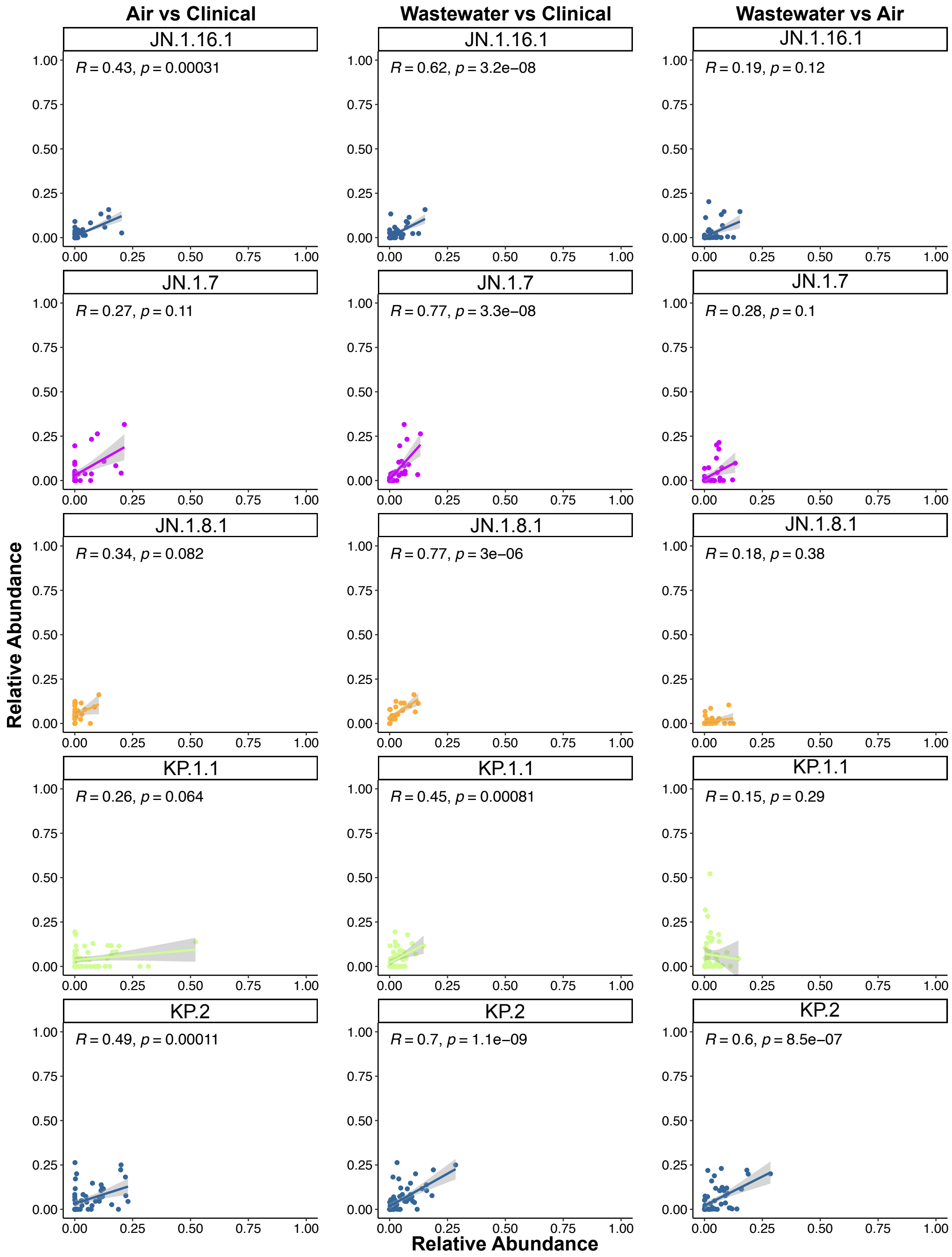

Air vs Clinical
Wastewater vs Clinical
Wastewater vs Air
JN.1.16.1
R = 0.43, p = 0.00031
R = 0.62, p = 3.2e−08
R = 0.19, p = 0.12
JN.1.7
R = 0.27, p = 0.11
R = 0.77, p = 3.3e−08
R = 0.28, p = 0.1
JN.1.8.1
R = 0.34, p = 0.082
R = 0.77, p = 3e−06
R = 0.18, p = 0.38
KP.1.1
R = 0.26, p = 0.064
R = 0.45, p = 0.00081
R = 0.15, p = 0.29
KP.2
R = 0.49, p = 0.00011
R = 0.7, p = 1.1e−09
R = 0.6, p = 8.5e−07
Relative Abundance
Relative Abundance

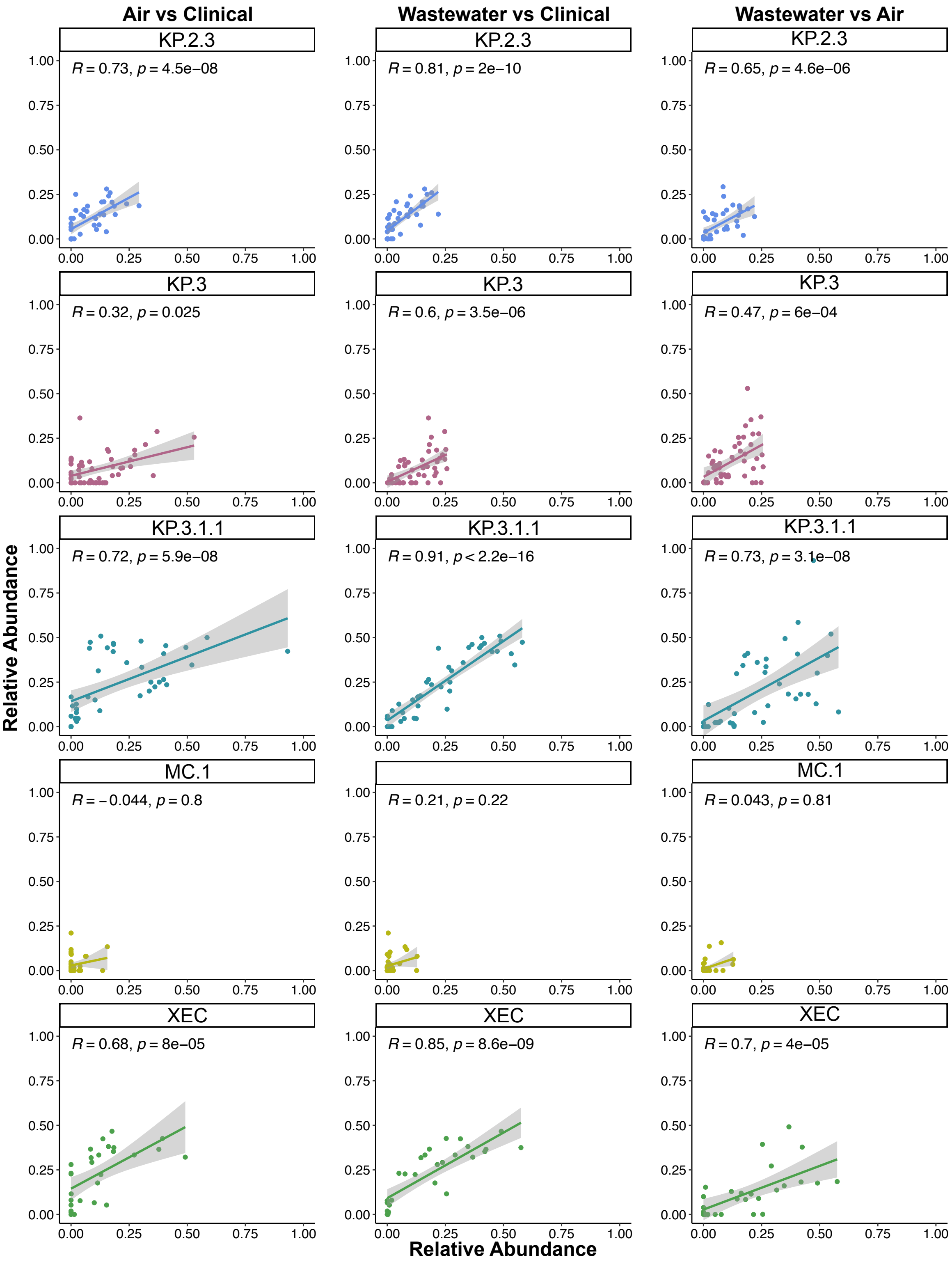

Air vs Clinical
Wastewater vs Clinical
Wastewater vs Air
KP.2.3
R = 0.73, p = 4.5e−08
R = 0.81, p = 2e−10
R = 0.65, p = 4.6e−06
KP.3
R = 0.32, p = 0.025
R = 0.6, p = 3.5e−06
R = 0.47, p = 6e−04
KP.3.1.1
R = 0.72, p = 5.9e−08
R = 0.91, p < 2.2e−16
R = 0.73, p = 3.1e−08
MC.1
R = −0.044, p = 0.8
R = 0.21, p = 0.22
R = 0.043, p = 0.81
XEC
R = 0.68, p = 8e−05
R = 0.85, p = 8.6e−09
R = 0.7, p = 4e−05
Relative Abundance
Relative Abundance
0.00
0.25
0.50
0.75
1.00

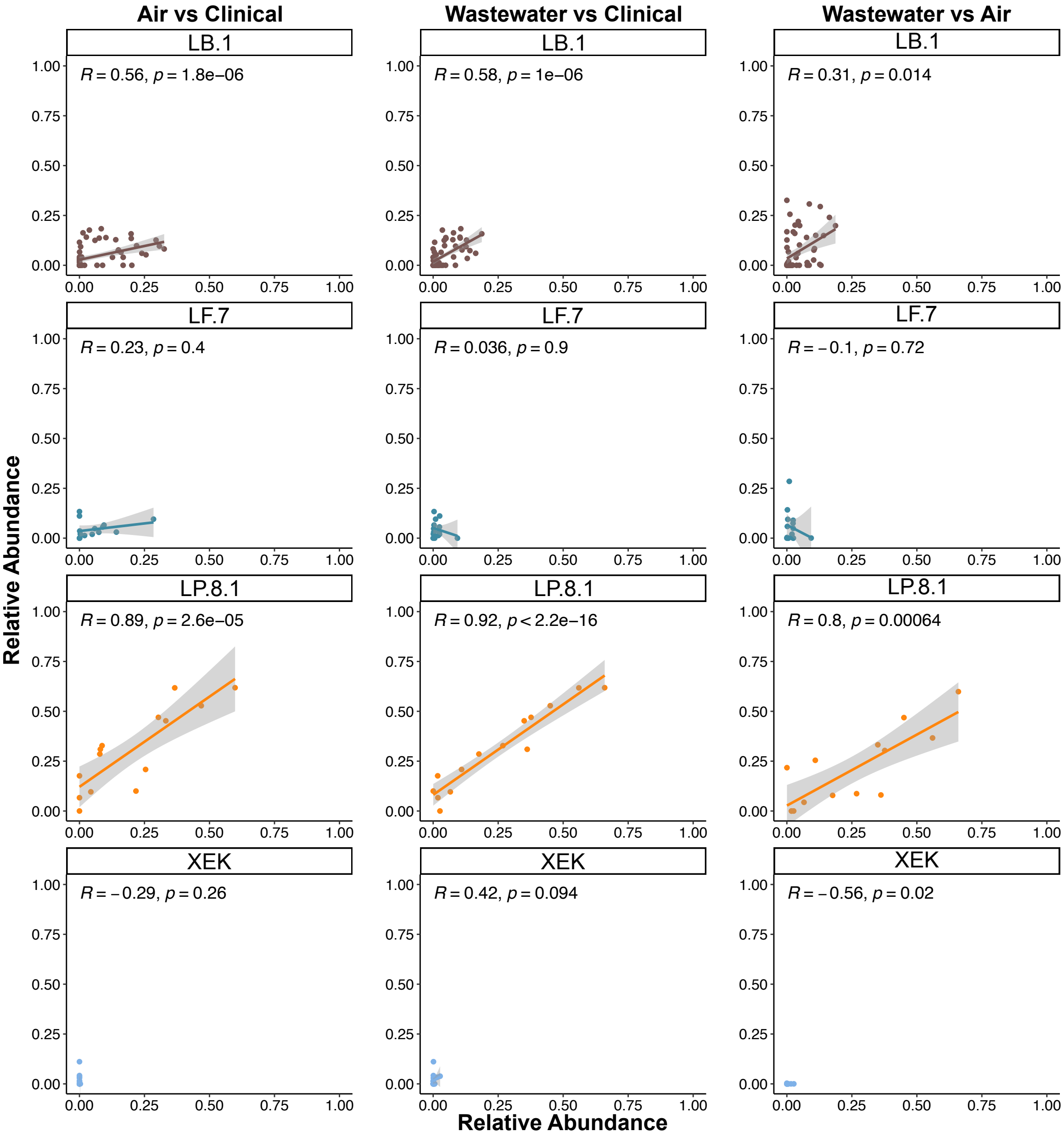


**Supplemental Figure 4.** Correlation plots of the relative abundance of all tracked variants in air vs clinical samples, wastewater vs clinical samples, and wastewater vs air samples. Linear regression line for each variant and 95% confidence interval (shaded) shown. Strength and significance of correlation was determined by Spearman correlations.

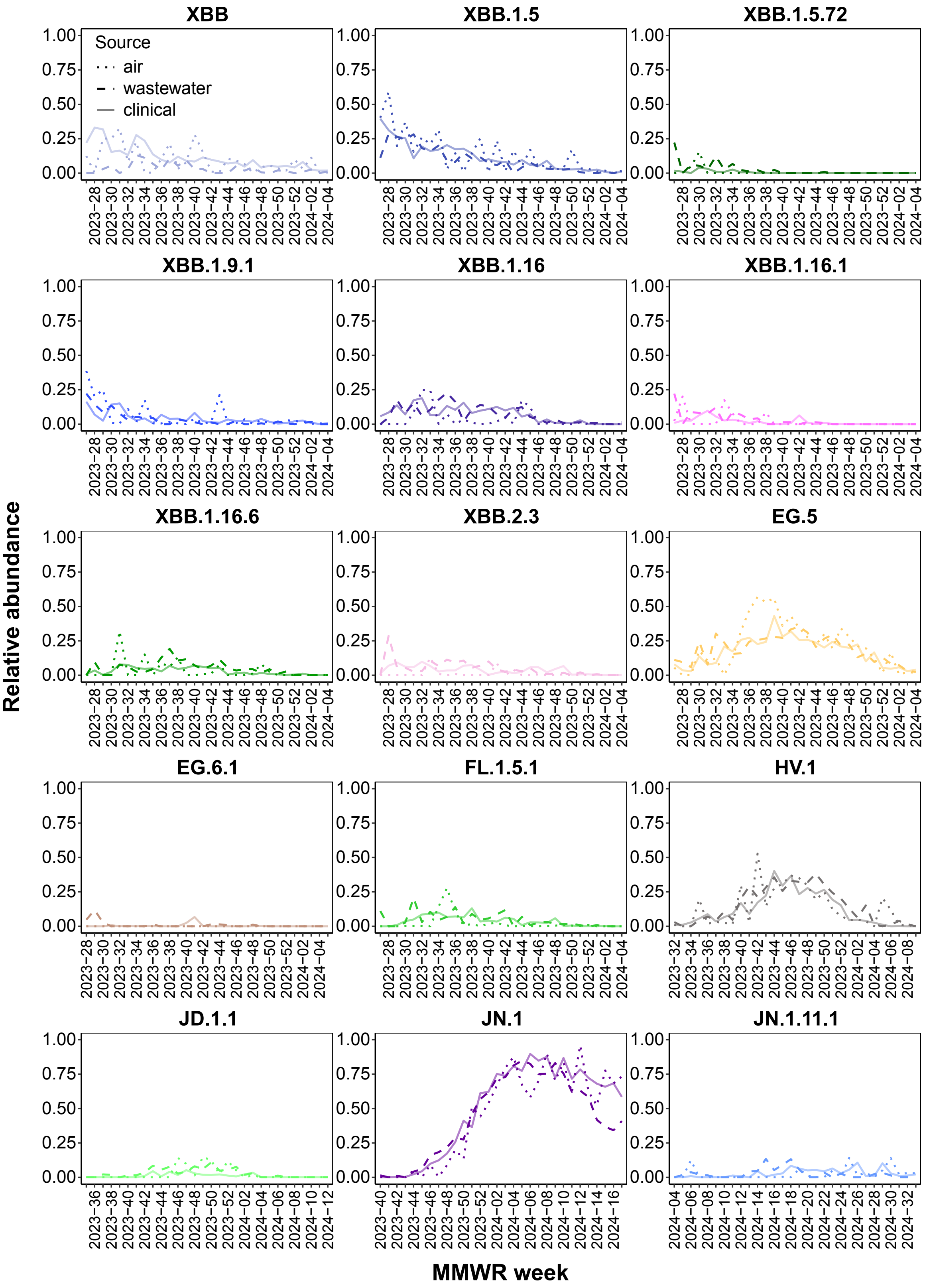
XBB
XBB.1.5
XBB.1.5.72
XBB.1.9.1
XBB.1.16
XBB.1.16.1
XBB.1.16.6
XBB.2.3
EG.5
EG.6.1
FL.1.5.1
HV.1
JD.1.1
JN.1
JN.1.11.1
Source
air
wastewater
clinical
Relative abundance
MMWR week

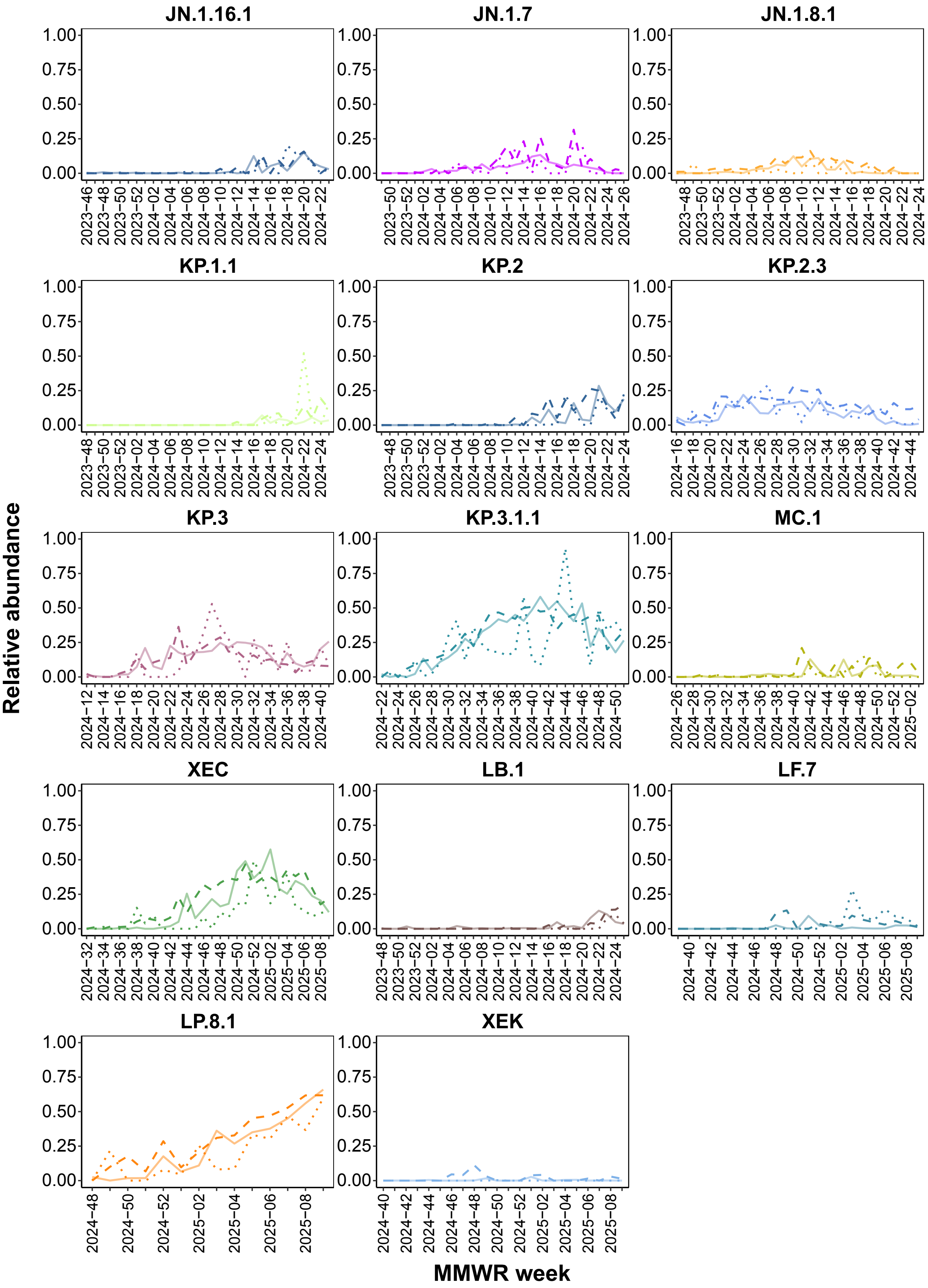
JN.1.16.1
JN.1.7
JN.1.8.1
KP.1.1
KP.2
KP.2.3
KP.3
KP.3.1.1
MC.1
XEC
LB.1
LF.7
LP.8.1
XEK
Relative abundance
MMWR week

**Supplemental Figure 5.** Relative abundance emergence of all 29 tracked variants in air (dotted lines), wastewater (dashed lines), and clinical (solid lines) samples. Up to the first 30 weeks after first detection and lineage designation (whichever came last) shown.

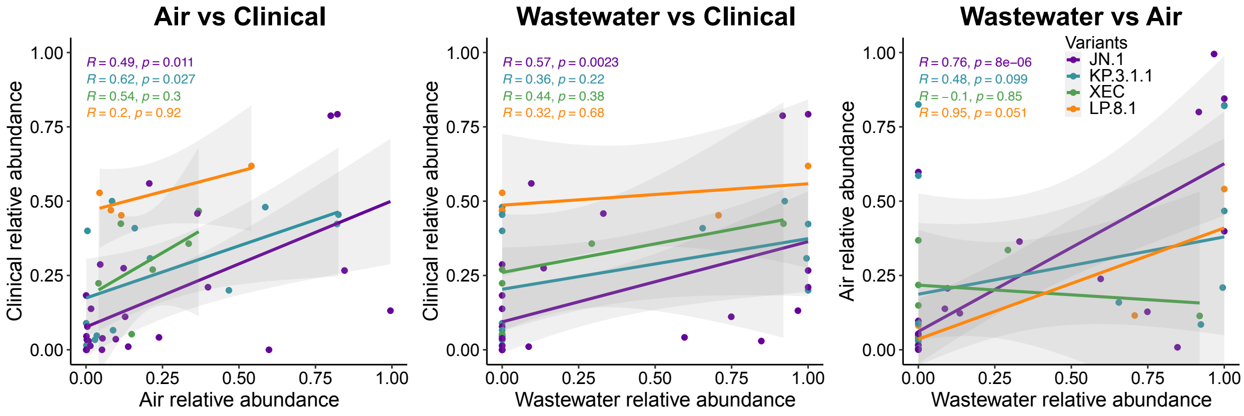


**Supplemental Figure 6**. Correlation plots of the relative abundance of four common variants in air vs clinical samples, wastewater vs clinical samples, and wastewater vs air samples from a single site. Linear regression line for each variant and 95% confidence interval (shaded) shown. Strength and significance of correlation was determined by Spearman correlations.

### Supplemental Tables

**Supplemental Table 1. Timing of respiratory peak and onset for air and wastewater compared to clinical surveillance**

| Pathogen | Dataset | Onset (2023-2024)[a] | Peak (2023-2024) | Onset (2024-2025) | Peak (2024-2025) |
|---|---|---|---|---|---|
| Influenza A | Clinical | 10-07-23 | 12-23-23 | 10-26-24 | 01-25-25 |
| | Air (all)[b] | +1 | +3 | +6 | -2 |
| | Air (ED)[c] | +1 | +3 | +6 | -1 |
| | WW[d] | +7 | +2 | +7 | 1 |
| Influenza B | Clinical | 11-11-23 | 03-09-24 | NA[e] | NA |
| | Air (all) | -5 | +1 | NA | NA |
| | Air (ED) | -5 | +1 | NA | NA |
| | WW | +2 | +2 | NA | NA |
| RSV | Clinical | 09-16-23 | 11-25-23 | 10-19-24 | 12-21-24 |
| | Air (all) | +4 | +7 | +7 | +7 |
| | Air (ED) | -2 | -4 | +5 | 0 |
| | WW | +2 | +1 | +4 | +3 |
| SARS-CoV-2 | Clinical | NA[f] | 12-23-23 | 06-08-24 | 08-17-24 |
| | Air (all) | NA | +8 | +8 | 0 |
| | Air (ED) | NA | -3 | +5 | -1 |
| | WW | NA | -2 | NA[f] | +4 |

[a]Difference in weeks compared to clinical data date

[b]Data from all air samplers

[c]Data from air samplers placed in emergency departments

[d]Wastewater

[e]Not applicable because influenza B peak was not captured by the end of the study period

[f]Not applicable because data never exceeded the threshold defined for onset

**Supplemental Table 2. First detection MMWR week for SARS-CoV-2 variants that emerged during the study period**

| Variant | Air | Wastewater | Clinical |
|---|---|---|---|
| JN.1.8.1 | 2023-49 | 2023-49 | 2023-47 |
| JN.1 | 2023-41 | 2023-42 | 2023-40 |
| KP.3 | 2024-13 | 2024-12 | 2024-12 |
| HV.1 | 2023-32 | 2023-32 | 2023-32 |
| KP.2.3 | 2024-16 | 2024-16 | 2024-16 |
| JN.1.11.1 | 2024-06 | 2024-04 | 2024-05 |
| XEC | 2024-34 | 2024-32 | 2024-33 |
| KP.3.1.1 | 2024-23 | 2024-22 | 2024-23 |
| LP.8.1 | 2024-49 | 2024-48 | 2024-49 |
| MC.1 | 2024-26 | 2024-28 | 2024-29 |
| JD.1.1 | 2023-46 | 2023-35 | 2023-37 |
| JN.1.7 | 2023-50 | 2023-49 | 2024-01 |
| LF.7 | 2024-39 | 2024-44 | 2024-48 |
| XEK | 2024-40 | 2024-41 | 2024-46 |
| KP.2 | 2023-50 | 2023-47 | 2024-04 |
| JN.1.16.1 | 2023-46 | 2023-46 | 2024-10 |
| XBB.2.3 | 2023-28 | 2023-27 | 2023-28 |
| KP.1.1 | 2024-10 | 2023-48 | 2024-14 |
| LB.1 | 2023-48 | 2023-48 | 2024-16 |
